# Mental health pathways and treatment as usual for young people experiencing depression in the United Kingdom: A mixed methods study

**DOI:** 10.64898/2025.12.03.25341537

**Authors:** Siobhan B. Mitchell, Vashti Berry, Juliette Westbrook, Emma Grace Carey, Alice Wickersham, Alice Garrood, Rachel Hayes, Julieta Galante, Husna Hassan Basri, Kristin Liabo, Tamsin Ford

## Abstract

**Background:** Epidemiological data indicates that the prevalence of depression among young people (YP) is increasing. Evidence-based interventions are effective for many, but 20 to 40% fail to respond and relapse is frequent and therefore, this study placed a particular focus on second-line treatments. The modification of psychological processes that predispose treatment failure or relapse necessitates the development of specific interventions for those who need them. But their evaluation requires clear understanding of current practice to compare and demonstrate effectiveness.

**Methods:** We explored treatment as usual (TAU) for young people (YP) receiving support for depression in a pilot 2-site feasibility trial comparing a Mindfulness for Adolescents and Carers (MAC) against TAU in Child & Adolescent Mental Health Services (CAMHS) in the UK. We collected qualitative data via interviews with senior managers and case-managing clinicians, as well as quantitative data via Treatment Recording Sheets completed by clinicians for participating YP and a clinical service audit. We synthesised these data across the two trial sites with the intention of providing a description of TAU that could be used to design a future definitive trial of MAC.

**Results:** While there were differences in the approach and provision between the sites, it was possible to produce an understanding of the main components and processes of CAMHS TAU for young people who have depression. Typical pathways include entry/referral, screening and intake; waiting/supportive space; first-line and second-line treatment/s; discharge and re-referral; and crisis support. The most common second-line provision was care co-ordination, provided to YP in both arms of the trial. YP participating in the pilot feasibility trial received different combinations of support and those allocated to MAC received less support from CAMHS than those allocated to TAU.

**Conclusions:** There is significant variability in the provision of services to YP experiencing depression. Future studies should compare the treatment plus care coordination with TAU.

## Background

Depression in children and young people is common, affecting between 1.7-3.9% of the population globally [1] with typical onset in the early teenage years [2]. In the UK, repeated cross sectional surveys indicate that young people’s mental health is deteriorating, with notable rises in emotional disorders especially among teenage girls [3, 4]. Despite national guidelines for evidence-based intervention [5], there is wide variability across the country in the funding allocated to resource treatments and interventions and hence in the services offered to children and young people [6]. Variation is not necessarily detrimental to outcomes – indeed it may reflect a system that is appropriately tailored to local context and needs – but it can point to inequalities in access while a lack of standardisation can produce difficulties for those evaluating the effectiveness of new interventions against current practice or treatment as usual (TAU). For example, in a recent network meta-analysis exploring the comparative efficacy of a range of treatments for depression in adolescents, the number and type of control conditions in included trials varied to the extent that the lack of a common comparator reduced network connectivity and analytical power in the conclusions [7].

Furthermore, while efficacy studies typically compare singular or ‘simple’ treatments against a defined control (placebo) or other active therapy, the choice and sequence of support offered to young people attending Child and Adolescent Mental Health Services (CAMHS) is determined by locally defined (clinical) ‘care pathways’. Care pathways are operational systems that translate clinical guidelines into processes for practice, allowing for mutual decision-making with the patient and, where relevant, integration with other health or social services. These pathways aim to standardise the management and co-ordination of care from the identification of initial difficulties through to discharge and maintenance support, and may be specific to a condition (e.g., depression) or a population (e.g., YP with learning difficulties) [8, 9]. This system-level variation makes the testing of treatment innovations additionally challenging because evaluators must both agree on an appropriate comparison/counterfactual against which to estimate their interventions’ impact *and* understand its placement in the care pathway across sites.

Current systematic reviews of clinical treatments for depression in children and young people [7, 10, 11] reveal a wide array of control conditions, including active controls, pill or psychological placebos, minimum attention control, wait-list, usual care or treatment as usual (TAU), web-based supportive contact, and medication management. Such varied comparators can lead to different estimated effects for the intervention being investigated, for example, with lower effects when compared with usual care versus wait-list [12]. Furthermore, where TAU is the comparator, standardising or protocolising care across study sites is uncommon and participants can be offered different types of care with different durations and intensity [13-17]. In other words, trials often assume sufficient similarity in provision across sites, arguably in line with best practice/treatment guidelines, and rarely report on the variability of TAU at individual or site level or across conditions. This reduces the interpretation of both effectiveness findings and intervention applicability/implementation in other contexts.

### Trialling Mindfulness-Based Cognitive Therapy

Mindfulness-Based Cognitive Therapy (MBCT) is established in the adult literature as effective relapse prevention for depression [18] but the evidence-base in children and young people is still in its infancy and it is not widely used in CAMHS [19]. Given its success in adults, we argue that exploring its potential for children and young people is important [19]. It is also highlighted as a research recommendation by NICE (see https://www.nice.org.uk/guidance/ng134/chapter/Recommendations-for-research).

The evidence for MBCT for adults is strongest for those with repeated depressive episodes, which suggests a possible role with YP as a second-line response for those with residual difficulty after a first-line evidence-based intervention and/or as a relapse and maintenance support after discharge [19]. Thus, appropriate comparators against which to evaluate the success of MBCT for YP are other second-line interventions in CAMHS depression care pathways, where they exist, but - to our knowledge - there have been no studies characterising TAU in mental health services for young people receiving secondary support for depression. Similar studies do exist for adult services [20]. Thus, in the context of designing a definitive trial of MBCT for YP, there is a need to understand both first and second-line treatments and whether this usual provision in different CAMHS sites across England is sufficiently similar. Given the increasing pressure of referrals and waiting lists for CAMHS, we need to assess whether participants allocated to receive the intervention receive similar TAU to those in the control condition.

### Research Question

This study aimed to answer the following research question: What are the current care pathways under CAMHS for adolescents experiencing depression (with or without anxiety) and what treatment/s is provided to them? We used mixed methods, combining qualitative interviews and quantitative data collected during the ATTEND Feasibility Trial with an audit of electronic health records at one of the two participating sites.

## Methods

### The ATTEND Feasibility Trial

TAU was explored in the context of the *ATTEND* feasibility pilot trial of a MBCT programme for young people and their carers, which ran from October 2020 to January 2022. The adapted MBCT programme – called Mindfulness for Adolescents & Carers (MAC) – consisted of 9 weekly group-based sessions, each lasting 1.5 hours, delivered to YP and, in parallel to their parents/carers (PC). Although initially designed as an “in person” intervention, due to Covid-19, sessions were attended via video-conference. Facilitators were either experienced mindfulness practitioners or CAMHS clinicians who underwent training prior to delivery. Multi-centre ethical approval was received from the East of England–Cambridge South Research Ethics Committee (ref: 20/EE/0246) and local research governance approval. Clinical trial number: not applicable.

Participants were eligible for the MAC intervention if they were aged 14-17 years, had completed one evidence-based, first-line treatment for depression but were not sufficiently well for discharge or had been rapidly re-referred. YP were excluded from the study if they required active clinical management for an eating disorder, post-traumatic stress disorder, psychosis, or self-harming behaviour. YP and their PCs were recruited from CAMHS and randomised on a 2:1 ratio to receive MAC intervention plus TAU or TAU alone. As figure 1 illustrates, 28 YP were consented and recruited into the study in two sites (London/Devon); 19 were randomised to MAC and nine to TAU. An associated 17 PC consented to participate in the MAC arm, and 6 PC in the TAU arm.

**Figure 1:**
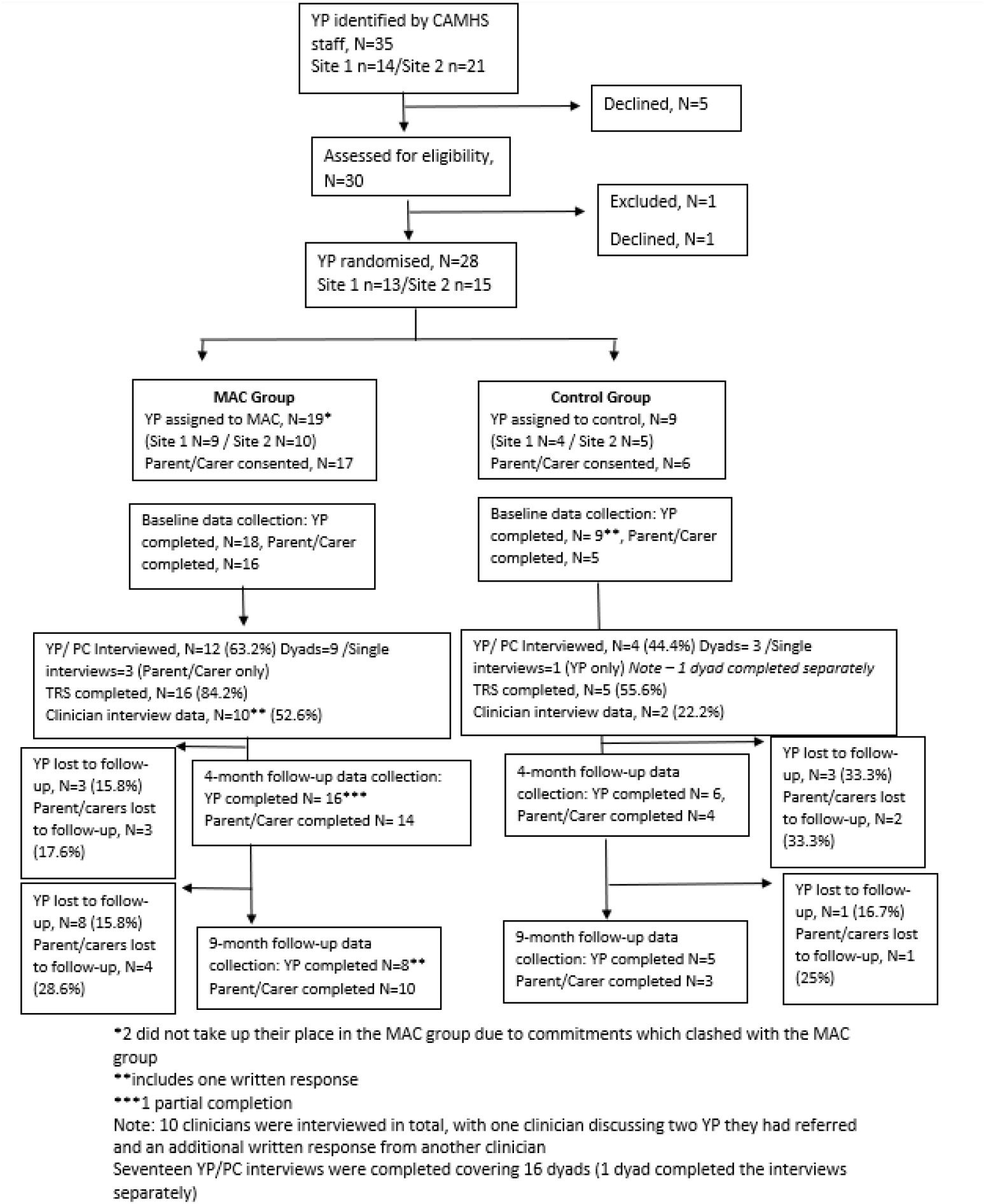
Flowchart of the ATTEND Feasibility Trial.

### Data collection

We gathered qualitative data from interviews with CAMHS managers and case-managing practitioners as well as quantitative data from a service audit and via a Treatment Recording Sheet [21] completed by CAMHS professionals for all YP in the study (See Figure 2). Interviews were carried out at the start of the study with 10 site-nominated, senior CAMHS managers across the two sites to gather accounts of care pathways in their service. Interviews were semi-structured (See Topic Guides in Appendix A) and lasted for approximately 30 minutes. All case-managing clinicians of YP involved in the trial were invited for interview at treatment completion. Interviews were conducted online between June and August 2021. The interviews were audio-recorded, transcribed verbatim and pseudonymised.

**Figure 2.**
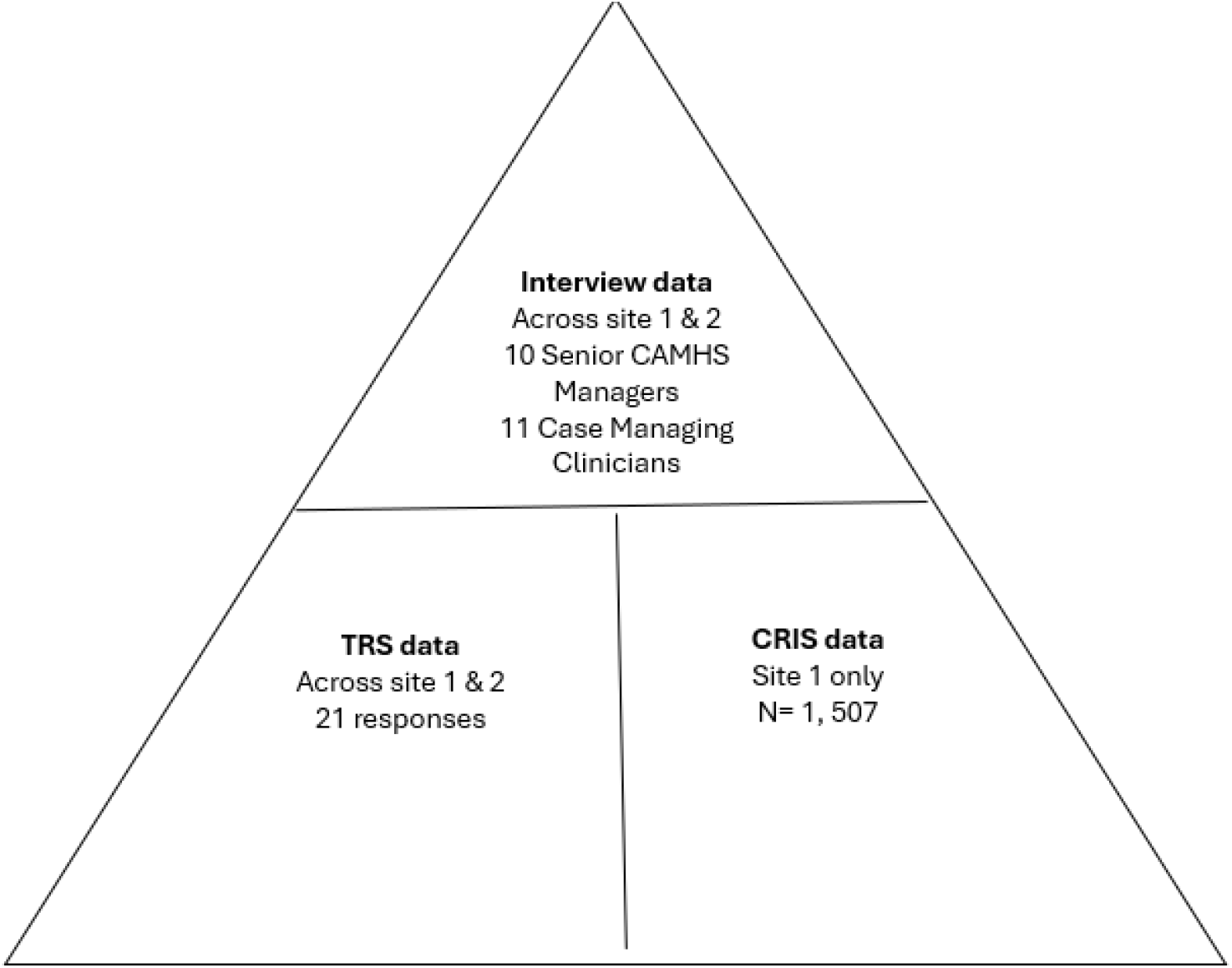
Summary of data collected.

#### The Treatment Recording Sheet

The Treatment Recording Sheet (TRS) (see Appendix B) captures components related to NICE-recommended approaches, including Interpersonal Therapy (communication skills training), Family Therapy (family engagement) and components of Brief Psychological Intervention (activity scheduling, psychoeducation, goal setting). The TRS captures which person each component has been delivered to e.g., YP, PC, family, school, and the frequency with which they were used on the following scale; frequent use (3), moderate use (2) and little use (1). YPs case-managing clinicians were asked at the 4-month follow-up to complete a TRS for therapy or interventions conducted over the past four weeks. By trial criteria, all young people involved in this study had already undergone a first-line treatment for depression, and therefore information captured on the TRS related to treatments and support offered as second-line.

### Analyses

Directed content analysis [22] was used was used to deductively code and analyse the interviews with senior managers and case-managing clinicians, to identify common and distinct service pathways and treatment experiences across sites as well as clinicians’ perspectives on using the Treatment Recording Sheet. Interviews were coded using NVivo 12 and pathways and treatments were extracted to generate a figure of components in both sites which were then synthesised. The figures were sense-checked by interviewees and revised accordingly.

Analysis of the TRS was undertaken using Stata 17 software. Numbers and percentages were used to summarise the distribution of the variables. The Mann-Whitney test was used to compare ordinal variables, and the Chi-squared test was used to compare binary variables between the trial arms. Data were then triangulated with the content analysis of interviews to assess the findings against NICE best-practice recommendations.

### Audit

At site 1, de-identified electronic health records can be accessed for research via Clinical Record Interactive Search (CRIS) [23]. Focusing on the window 1^st^ January 2018 to 31^st^ December 2019, we identified all children and adolescents who met the following inclusion criteria: (i) had an active CAMHS referral during the window (henceforth referred to as the ‘index referral’), (ii) the index referral was accepted when the patient was aged 14 to 17 years, and (iii) the patient received a depression diagnosis during the index referral (ICD-10 codes F32.x, F33.x, F34.1, F38.x, F39.x or F41.2 in structured diagnosis fields).

A total of n=1507 patients were eligible for inclusion. We summarised demographic and clinical characteristics of these referrals, and details of the treatment pathway, using a combination of structured fields and natural language processing (NLP). NLP is an automated method of extracting information from large volumes of text, and in this study was used to identify mentions of therapy episodes and psychotropic medication.

CRIS has received research ethics committee approval as a database for secondary analysis (Oxford REC C reference 18/SC/0372), and aspects of the audit described here were approved by the CRIS Oversight Committee. CRIS operates on an opt-out basis.

### Synthesis

Data were narratively and visually (in the form of a pathway diagram) synthesised across the methods detailed, to produce an understanding of care pathways and treatment as usual for young people with depression in CAMHS. Findings were triangulated to identify convergent and contradictory messages [24, 25]. A pathway diagram was created from the service component tables and shared with clinicians and service managers in both sites to sense-check its relevance to their site/service. Finally, multiple discussions were held between members of the ATTEND study team to explore the interpretation of the findings and to agree a conclusion regarding future trial design.

### Patient and public involvement

During the design phase of the study, researchers worked with a group of ten parents, carers and young people who had gone through CAMHS and attended group-based mindfulness sessions as a second-line treatment in the past. Once funding was secured, they formed a study involvement group. This group helped shape the study information materials, the recruitment process, data collection forms and the interview questions. We also met with two separate groups of CAMHS-experienced young people. One parent participated in the monthly study team meetings.

## Results

Interviews were carried out at the start of the study with 10 site-nominated, senior CAMHS managers across the two sites to gather accounts of care pathways in their service. Ten case-managing clinicians consented to be interviewed and reported on services provided to 10 (52%) MAC participants and 2 (22%) YP allocated to TAU. TRS responses were received for 21 YP participants’ care (21/28; 75%) from 18 case-managing clinicians.

**Table 1:** Breakdown of the location and trial arm of TRS data.

|  | MAC Arm | TAU Arm | Total |
| --- | --- | --- | --- |
| Site 1 (London) | 8 | 2 | 10 |
| Site 2 (Devon) | 8 | 3 | 11 |
| Total | 16 | 5 | 21 |

Of the patients included in the audit who had available gender data, most were female (n=1076, 71.6%), followed by male (n=410, 27.3%) and other genders (n=17, 1.1%). The mean age at index referral acceptance was 15.4 years (SD 1.09), and the mean age at depression diagnosis was 15.7 years (SD 1.09).

No statistically significant differences were found between the trial arms, with the exception of mental health interventions with the child, which was used more frequently in the TAU trial arm (P=0.03), However, given the small sample size, there was little power for this analysis, so we cannot conclusively rule out the possibility of other meaningful differences. Full descriptive statistics and p values from comparisons between trial arms can be found in the supplementary materials.

In the audit, most patients (n=1002, 66.5%) had at some point received another mental health diagnosis other than depression (defined as any ICD-10 F code other than those used to define depression in this audit). In total, n=530 (35.2%) had ever received an anxiety diagnosis (ICD-10 F4 codes other than F42.1 mixed anxiety and depression), n=161 (10.7%) an eating disorder (F50 codes), n=137 (9.1%) a personality disorder (F60/61 codes), n=222 (14.7%) a learning/neurodevelopmental disorder (F7/8 codes), n=60 (4.0%) a hyperkinetic disorder (F90 codes), and n=32 (2.1%) a schizotypal disorder (F2 codes).

Figure 3 is a visual representation of the synthesis of care pathways across the two sites, indicating the contact/s and intervention/s offered to YP from entry to discharge (including re-referral). The sections that follow provide a narrative of each component of the pathway.

**Figure 3.**
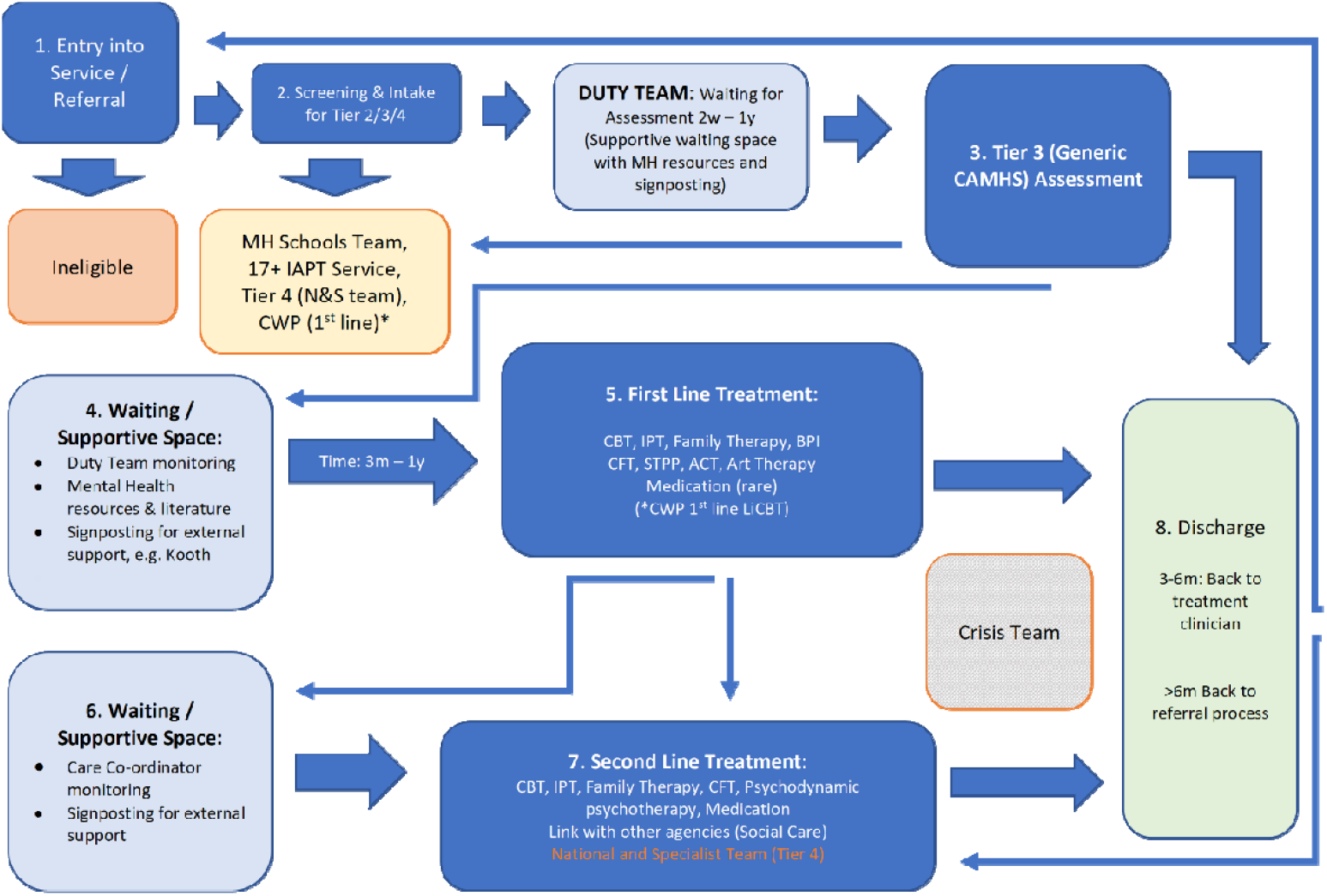
Visual representation of synthesis of care pathways across two sites. **Terms:** Mental Health in Schools Team (MH Schools Team) / Improving Access to Psychological Therapies Service (IAPT Service) / National and Specialist Team (N&S Team) / Children’s Wellbeing Practitioner (CWP) / Cognitive Behavioural Therapy (CBT) / Interpersonal Psychotherapy (IPT) / Brief Psychological Intervention (BPI) / Compassion-focused Therapy (CFT) / Short-term Psychodynamic Psychotherapy (STPP) / Acceptance and Commitment Therapy (ACT) / Low Intensity Cognitive Behavioural Therapy (LiCBT)

### (1) Entry/Referral – Screening and Intake

In the audit, the most common referral source was general practice (n=599, 39.7%), with smaller numbers referred from A&E (n=232, 15.4%), education settings (n=123, 8.2%), social services (n=54, 3.6%), and self- or carer-referrals (n=36, 2.4%). The remainder were referred from a wide range of other sources, including other health services.

Interviews with CAMHS managers revealed that, following referral, young people undergo screening for eligibility. Initial assessment described by clinicians varied across the two sites but encompassed the same core components. Site 1 clinicians described a full CAMHS assessment involving review of presenting difficulties and history of difficulties, developmental and family history, educational history, family composition and current family circumstances, family history of mental illness, and risk to self/others/from others. Site 2 clinicians described a choice assessment as part of the Choice and Partnership Approach (CAPA) approach [26]. This involved a similar review of those aspects noted by site 1 clinicians with an emphasis on discussion with the YP and parents about what they want, and what they think might work. Both sites mentioned use of standard measures, including Revised Child Anxiety and Depression Scale (RCADS) and Strengths and Difficulties Questionnaire (SDQ) as the national standard and others based on clinician discretion [27]. YP assessed as not meeting the threshold for specialist CAMHS are re-routed to Mental Health Support Teams (MHST) in schools, adult Improving Access to Psychological Therapies (IAPT) services or to a Child Wellbeing Practitioner (CWP), who offer lower intensity interventions. If, during screening, young people exceed the threshold for specialist CAMHS, they can be referred directly to a tertiary service. YP deemed eligible for specialist CAMHS support are seen by the Duty Team for assessment. Waiting times for assessment vary considerably and CAMHS managers suggested this can take anywhere from 2 weeks to one year, depending on the service/team/type of assessment. In the audit, of the n=1435 who attended a face to face/virtual appointment at some point during their index referral, the time between referral acceptance and first attended face to face appointment ranged from less than 1 day to over 2 years, with a median of 32 days (IQR: 7 to 98). This aligns with interviews with senior and case-managing clinicians who described a ‘crisis pathway’ where ‘RAG ratings’ (red, amber, green) are used to prioritise those presenting to the service in crisis. Similarly, if a young person is re-referred into the service for a further depressive episode, the intake process can differ to first entry (as discussed below).

Following assessment, young people are directed to the respective waiting list for the selected first-line treatments.

### (2) Waiting/Supportive Space

Waiting times for treatment varied significantly, often due to high caseloads and the availability of clinicians with particularly skills or expertise in specific interventions: *“there’s not as many people that do it [IPTA], so it’s a longer wait” A2107LT_case-managing clinician interview*.

Senior managers and clinicians indicated that prior to assessment and/or while waiting for first-line treatment, young people may receive a variety of mental health resources and literature, as well as signposting for external support (e.g. ‘Kooth.com’). During this period, it is the responsibility of the Duty Team - clinicians on call during working hours – to support YP if urgent needs arise.

As explained below, YP with residual symptoms after receiving a first-line treatment are directed for follow-on or more specialised second-line treatment and are often placed in a further waiting/supportive space until this support is available. Interviews with case-managing clinicians indicated that many young people receive ‘care co-ordination’ during this waiting period. This involves review meetings with a consistent, named clinician who monitors the YP’s mental state and risk status. It may also involve low-intensity psychoeducation. Despite the best intentions to provide ‘the right treatment at the right time’, young people are often matched with clinicians who have time/space, regardless of whether their skillset is appropriate for the YP’s needs: *“I was the only one that had any space. So*… *it’s as simple as that, really. She wasn’t given to me because I was more suited; it was just that I literally had a space*.*”* X21VC*_case-managing clinician interview*. Furthermore, the interviews with case-managing clinicians revealed that the skills or principles of psychoeducation utilised were often similar to those used during first-line treatment, which had not successfully alleviated symptoms or changing maladaptive thinking patterns if the young person required further intervention. Interview data revealed that clinicians did not always consider care co-ordination to be an appropriate or sufficient response and that often, referral on to a second line treatment was unsuccessful:

> *I’ve referred [to a second line intervention] and they’re saying she doesn’t meet the criteria so then I suggested perhaps low level CYP work in terms of addressing her panic attacks and low mood and, yeah, I referred back in April, I think, and we’re still waiting and every week I’m doing basically all the work and checking in with her. Every week she’s contacting me and saying, “I’m not too well. I’m not too good*,*” and I’m having to keep working with her. So, for an entire year that’s what I’ve been doing. So there’s a massive gap in what we say we can offer and what we actually offer” …*.*” It’s a kind of mix match of whatever’s available and sometimes it’s inappropriate. That’s where CAMHS is at the moment with their interventions. I don’t think we have a robust enough process to get young people to the right places at the right time in a seamless way, at the moment*.*”*
>
> *X20CM case-managing clinician interview*
>
> *…it feels like a quite substantial of time that I’ve been working with this young person. So… which I think is quite rare sometimes that we’re able to continue that continuity. But at the same time, within that pathway I know that some young people can get quite stuck, and I think having something new like the MAC study to try and give that a different spin on things was definitely helpful for my young person to try and prevent her from just waiting for an intervention” “I wasn’t doing any therapeutic work with her, but I was just having a well-being conversation and checking in to make sure she was okay, and there was no side-effects of the medication, and those sorts of things. So, I was kind of just, like I said, not doing any therapeutic work but just checking in and making sure that the risk was staying stable while she was doing that, and that she wasn’t having any side-effects from the medication whilst it was being increased*.
>
> *X2106SM case-managing clinician interview*

### (3) First-line Treatment

Interviews with the senior managers suggested that first-line interventions offered to YP are wide-ranging, dependent on their needs and the skills of the clinician/s available to support them, including: Cognitive Behavioural Therapy (CBT); Interpersonal Psychotherapy (IPT); Family therapy, Brief Psychosocial Intervention (BPI); Compassion-Focused Therapy (CFT); Short-Term Psychodynamic Psychotherapy (STPP); Acceptance and Commitment Therapy (ACT); Art Therapy; and medication.

This is supported by data from the audit, where we used a tool to identify ‘therapy episodes’ as a series of consecutively attended appointments identified as talking therapy sessions through a combination of event data and NLP [28]. Using this method, at least one therapy episode was identified during the index referral of n=333 (22.1%) included patients. Some patients underwent multiple therapy episodes during the index referral. Focusing on the first such therapy episode for each patient, the median number of sessions attended was 8 (IQR: 5 to 17). Where modality could be identified (n=31), first therapy episodes and was most frequently CBT (n=12). However, interviews with case-managing clinicians suggested that the treatment offered to YP generally consisted of either general talking therapy, CBT or systemic family therapy in the first instance. The duration of first line treatment was described in interviews as wide ranging but usually between six to eight sessions fortnightly over 12-14 weeks depending on treatment modality:

> *The partnership would normally. Normally be around 6 to 8 sessions, but there is the scope to make it longer if necessary, but usually if it’s going to be much longer, the idea is that you might need to move it into specialist, but that could be with the same clinician*.
>
> *C1X_Senior Clinician interview*

RAG (red, amber, green) ratings were also described, whereby clinicians use these to prioritise cases which come into the service in relation to level of risk. Interviews with senior managers and case-managing clinicians described widespread use of supportive treatment and generic partnership work. In Site 2 (Devon) this was largely influenced by following the Choice And Partnership Approach (CAPA) where a collaborative initial partnership is used to engage clients, followed by referral onto a more specific therapeutic treatment if needed: *“*…*they should have their choice appointment and some generic partnership. And then if they need something specific they come into a pathway or family therapy or CBT or whatever” C4X_Senior clinician interview*. This was supported by data from the TRS, which demonstrated moderate to frequent use of ‘Supportive Listening’, defined as ‘Reflective discussion with the client to demonstrate warmth, empathy, and positive regard without suggesting solutions or alternative interpretations’ across both arms.

If first-line treatments result in sufficient symptom reduction, the YP is discharged and/or signposted for less intensive community support: *“… we would have signposted to some kind of online services, counselling service that we use, and then there would have been the option to re-refer if they needed more support within our service in the future*.*” A2108KP_case-managing clinician interview*. Clinicians described using a combination of outcome measures such as RCADS and clinical judgement when making decisions about discharge of CYP from the service:

> *So I think the outcome measures, the parent reports, the young person reports would all inform the clinical judgement, but in the end it’s a clinical judgement and not just about symptoms, but about functioning. So if we’re lucky, we see kids who are, I don’t know, back in school, back in college, engaging with friends. The functioning is then a real indicator that things have changed*
>
> *C7A_ Senior Clinician interview*

However, if sufficient progress is not achieved, or depressive symptoms are still present following the completion of first-line treatment, young people are referred on for a second-line treatment.

### (4) Second-line Treatment

Interviews with CAMHS managers suggested that second-line treatments included CBT, IPT, Family Therapy, CFT, and Psychodynamic Psychotherapy (PP) and/or medication. Where required, young people would also be linked to other agencies to attend to wider psychosocial needs, e.g. social care or national / regional highly specialist teams.

The case-managing clinician interviews, and audit suggest that medication use was common across both sites, with the TRS indicating that antidepressant medication was used by n= 7/21 (33%) participants. In the audit, we used an NLP tool to detect mentions of psychotropic medications during the index referral. In total, n=736 (48.8%) included a mention of an antidepressant, n=469 (31.1%) a sleeping medication/tranquiliser, n=240 (15.9%) an antipsychotic, and n=54 (3.6%) a mood stabiliser.

### (5) Discharge and re-referral

Interviews with senior clinicians suggested that young people can be discharged at multiple time points (see Figure 3) including following the completion of a first- or second-line treatment depending on symptom severity.

Of the sample of young people involved in the ATTEND trial (n=28), at the 9-month follow up, 46.4% had not been discharged and remained in CAMHS. In the audit, n=1439 (95.5%) had been discharged by the time of data extraction. Of these, two thirds were discharged back to their GP (n=962, 66.9%). Most were discharged on professional advice or with clinical consent (n=1290, 89.6%), while the remainder were discharged against professional advice or due to non-attendance (n=114, 7.9%), or were redirected without engagement (n=35, 2.4%). Among those discharged, the median time spent in the service from referral acceptance to discharge was 421 days (IQR: 194 to 777).

Entry into the system via re-referral (e.g. for further related episodes) differs depending on the time since discharge. Clinicians at both sites indicated that if this is within 3-6 months of discharge, the YP would be referred directly back to the most recent treatment clinician. If referred after a period of more than 6 months, they would go back to the beginning of the care pathway process (i.e. entry, screening and intake): *“…we have a three-month time frame if they feel like actually things are deteriorating or they feel like they need additional support, they kind of come back in and go back onto the treatment waiting list rather than the assessment waiting list*.*” A2107LT_case-managing clinician interview*.

There were exceptions to this rule, based on previous experience with the service or relationships with practitioners:

> *…because I’d worked with this young person previously a number of years before, and because of the area they lived in, they actually came straight to me, so I was immediately able to offer a CBT intervention to them without them having to go through the core partnership sort of side of CAMHS, as it were*.
>
> *X2107BJ_case-managing clinician interview*

Of the n=1507 included in audit analysis, n=692 (45.9%) had had another referral to the Trust’s mental health services (either accepted or rejected) prior to the index referral. Of those discharged, n=632 (43.9%) had a subsequent referral to the Trust’s mental health services (either accepted or rejected) by the time of data extraction. If a case is re-opened due to re-referral within a limited timeframe post-discharge, it could appear as part of the index referral rather than a subsequent referral.

Of the n=1507 index referrals included in the audit, n=945 (62.7%) were still active when the patient turned 18 years old, and so may have transitioned or been eligible to transition to adult mental health services.

### (6) Crisis and Risk

Symptom severity influences waiting times, the teams that YP access and the treatments offered. Referrals to CAMHS teams can be classed as ‘routine’, ‘urgent’ or terms relating to the referral being more urgent/an emergency. If YP are experiencing a mental health crisis or are referred to the psychiatric liaison team through A&E, assessments are administered immediately.

In the audit, a small proportion of index referrals were marked ‘urgent’ or ‘within 7 days’ (n=139, 9.2%), and the remainder were marked ‘normal’ or ‘within 28 days’ (n=1365, 90.8%). Referral urgency was associated with time to first contact. Of the n=1435 who attended a face to face/virtual appointment at some point during their referral, the median time from referral acceptance and the first such contact was 0 days (IQR: 0 to 1) when the referral was marked ‘urgent’ or ‘within 7 days’, as compared to 38 days (IQR: 12 to 111) when the referral was marked ‘normal’ or ‘within 28 days’. Only a very small minority were marked ‘within 28 days’, so the median of 38 days may not indicate that this time limit was exceeded for those patients, as the median would be influenced by the rest of the sample. Similarly, the median time from referral acceptance and the first attended face to face/virtual contact was 0 days (IQR: 0 to 1) when A&E was the referral source, compared to 42 days (IQR: 16 to 117) for other referral sources.

The clinician interviews informed us that if a referral is urgent, these are reviewed within 24-48 hours. Interviews with senior clinicians working in the crisis teams described how RAG ratings are used to prioritise individuals who present to the service in crisis:

> …*So obviously came in as a crisis, in the crisis pathway, and then was immediately transferred over to*… *we don’t have specific pathways in our team, so she was transferred onto what we call the partnership*…*Waiting list as a high-risk amber case. So she would have been prioritised for treatment, and I was the only one that had any space*.
>
> *X2107BJ_case-managing clinician interview*

Overall risk was captured in the TRS, with critical incidents including deliberate self-harm, and attempted suicide indicated in 8/21 (38%) of the forms.

## Discussion

We aimed to explore the TAU in CAMHS for YP with depression, through the lens of clinician reports, interviews and audit data. Our findings offer valuable insights into the complexities and challenges of providing mental health care within CAMHS. The study examined the services provided to young people in two distinct sites during a feasibility trial and documented the broad range of intervention and support on offer throughout the treatment pathway. Although there was considerable variation in the types and extent of interventions and support provided across the two study sites, we identified a common overarching care pathway for treating depression in both locations. This pathway included key stages such as entry and referral, screening and intake, waiting and supportive space, first-line and second-line treatments, discharge and re-referral, and crisis support.

### Clinical Pathways and Referrals

Our analyses indicate a high reliance on primary care (GPs) and Accident and Emergency (A&E) departments as referral sources for YP, which aligns with previous studies suggesting these settings are critical in initiating mental healthcare for adolescents [29]. However, this also underscores a potential gap in earlier detection and intervention for YP at the community or school level [30, 31]. Clinicians noted a significant variation in the referral process, especially in terms of waiting times and access to appropriate clinicians, with wait times ranging from less than a day to over two years. This variability likely reflects the broader systemic pressures facing CAMHS, including staff shortages, high caseloads, and resource limitations [32].

Earlier feasibility work at Site 2 indicated a disconnect between policy recommendations, such as those from NICE [5], and the realities of service delivery [33]. This gap between ideal practice and real-world implementation was also evident in this study. Some aspects of NICE guidance, such as the emphasis on building a supportive and collaborative relationship (1.1.3), were commonly observed across the care pathways. Pile et al. (2019) also reported “mostly good” adherence to NICE guidelines within one Trust they audited [34].

As anticipated, the YP who participated in this feasibility trial received varying combinations of support from the service providers. Those in the intervention arm, who received MAC during the study period, generally had fewer contacts with services and did not receive other interventions compared to those assigned to TAU. It is conceptually important when planning a randomised controlled trial, to clarify whether the study will test the intervention against or in combination with TAU. Our data support the intuitively plausible response of reduced support offered to YP receiving MAC as part of a research trial, due to the practitioner perception of support and less help seeking by the YP, which suggests that actually this is a false dichotomy.

A key component of TAU across both arms of the study was care coordination, where a designated professional oversees the care plan and advocates for the young person. This suggests that care coordination could be pre-specified as a standard component in future trials to enhance comparability. However, while care coordination is an important function, it is not in itself a direct form of treatment but rather an administrative role that requires time and resources. Despite being a common feature in this study, the extent to which care coordination truly delivers “care” or effects therapeutic change is limited. If no other forms of treatment are being provided, the role of care coordination in delivering meaningful care may need further exploration, as the clinician time and resource devoted to it might be more effectively deployed in active treatment.

### Waiting and Supportive Care

Both the clinician interviews and audit data suggest significant waiting times for first- and second-line treatments. The period spent waiting for treatment is a critical juncture in the care pathway, during which YP may face worsening symptoms [35, 36]. While some YP received supportive interventions such as psychoeducation and care coordination during this period, the interviews revealed concerns about their adequacy and appropriateness. Clinicians expressed frustration with the mismatch between the available resources and the complex needs of YP, with many describing instances where care coordination fell short of meeting the therapeutic needs of the individual. This echoes findings from other research indicating that prolonged waiting times and insufficient support during this period can exacerbate mental health issues, potentially leading to further distress or crises [37, 38].

Furthermore, the reliance on “whatever is available,” rather than tailoring interventions to specific needs, raises an issue around mismatched care, which was particularly evident when YP awaited specialized treatments after initial interventions had not yielded significant improvements. As clinicians reported, this gap in services is a significant challenge for both YP and service providers.

### First-line and Second-line Treatments

Our data also highlighted a wide range of first-line treatments offered to YP, which were often delivered in a flexible manner, depending on the clinician’s skill set and YPs specific needs. The use of CBT, in particular, was prominent, consistent with evidence supporting its efficacy in treating adolescent depression [5]. However, the relatively short duration of first-line treatments (typically 6-8 sessions) may not be sufficient for all YP, particularly those with more severe or complex presentations, and the need for second-line interventions was noted for those who did not respond to initial treatments [36].

Despite the availability of second-line treatments, such as CBT, IPT, and psychodynamic psychotherapy, clinicians expressed concerns about the adequacy of these services, particularly in cases where YP had residual symptoms following first-line treatment. In some instances, the referral process for second-line care was described as cumbersome, with YP experiencing delays due to restrictive eligibility criteria. This issue reflects a broader concern within mental health services, where resources are stretched thin, and access to more specialized care is often limited or delayed [3, 39].

### Discharge and Re-referral

Although most YP were discharged from CAMHS services, many still experienced residual symptoms and were re-referred into the system for further care. The re-referral process varied depending on the timing of re-entry into the system, with those re-referred within six months able to bypass the initial intake process. However, the need to re-enter the intake process after a longer break could contribute to further delays.

Moreover, the audit revealed a high rate of re-referral, with many YP requiring further support after their discharge, which may be indicative of insufficient initial treatment and insufficiently stringent discharge criteria in at least some cases. Repeated referrals also highlight the ongoing vulnerability of this group, with YP potentially falling through the cracks between different levels of care or experiencing disruptions in continuity when transitioning between services [40].

### Crisis and Risk Management

Risk management, particularly in cases of crisis, was another significant theme. The study revealed that the severity of symptoms, such as self-harm and suicidal ideation, played a central role in determining the urgency and type of care provided. In line with other studies, clinicians reported the use of a RAG rating system to prioritize cases, ensuring that those in immediate crisis received urgent attention, but which also emphasised the sheer volume of cases and the risks associated with delayed care [29]. Clinicians noted that despite attempts to prioritize based on risk, the availability of services and clinicians often meant that treatment was not always ideally matched to the needs of the young person.

### Strengths and Limitations

This study triangulated data from qualitative and quantitative approaches from two CAMHS sites in England, covering urban and semi-rural areas. While the variability between these sites is important, it is likely that there is significant variation in service provision across the country [41]. Thus, the care pathway identified in this study should be further explored across additional sites to assess its generalisability.

Another limitation is that our feasibility trial was conducted during the COVID-19 pandemic, which affected the delivery of MAC and will undoubtedly have similarly influenced TAU reported via the TRS. However, the audit used data from 2018-2019, prior to the pandemic, which produced similar findings to a similar audit of depression treatment across Cambridgeshire and Peterborough [41]. Given the well-documented impact of COVID-19 on CAMHS referrals, eligibility thresholds, and waiting times, our findings may not be fully applicable to the post-pandemic context [42].

Additionally, while the data on gender, age, and diagnoses provide valuable demographic context, further research is needed to explore the intersection of social, environmental, and psychological factors that may influence treatment outcomes for YP with depression.

## Conclusion

Our findings highlight the complexity of the care pathways for young people with depression within CAMHS. While a broad range of treatments are available, significant resource issues including long waiting times and mismatched care continue to hinder these services. There is considerable variability while care coordination was the most common second-line service provided to young people in both arms of the trial, but is not actively therapeutic. Future studies should investigate the comparative effectiveness of treatment plus care coordination versus treatment as usual.

## Supporting information

Supplementary Table 1. TRS

Appendix B TRS

Appendix A Topic Guides

## Data Availability

All data produced in the present study are available upon reasonable request to the authors

## Abbreviations

TAU: Treatment as Usual
CAMHS: Child and Adolescent Mental Health Services
MAC: Mindfulness for Adolescents and Carers
MBCT: Mindfulness-Based Cognitive Therapy
TRS: Treatment Recording Sheet
NICE: National Institute for Clinical Excellence
YP: Young People
PC: Parents/Carers

## Declarations

### Ethics approval and consent to participate

Multi-centre ethical approval was received from the East of England–Cambridge South Research Ethics Committee (ref: 20/EE/0246) and local research governance approval. The research was conducted in accordance with the Declaration of Helsinki.

### Consent for publication

Not applicable.

### Availability of data and materials

The data that support the findings of this study are available from the corresponding author on reasonable request.

### Authors’ contributions

TF, VB, RH, KL along with other study co-applicants devised the concept for the research and study design. KL coordinated PPIE input throughout the project. SM, JW, EC, JG & HHB collected general study data and conducted interviews. JW and AW conducted the audit. AG analysed TRS data. SM, JW & EC analysed interview data. VB, RH, JG supported with analysis and interpretation of data. VB, SM, AG completed synthesis of data. SM led on the writing of the manuscript, supported by AG, TF, VB, JW and AW. All authors read and approved the final manuscript.

## Acknowledgements

Fabiana Mariscotti, Maria Tejerina-Arreal and Hayley gains supported with the project. PPIE advisory groups: the group of young people and carers established specifically for this study, as well as two groups of CAMHS experienced young people.

