## Supplementary Table 1. TRS for "Mental health pathways and treatment as usual for young people experiencing depression in the United Kingdom: A mixed methods study"

| Outcome (Person)  Frequency of use (0-3) | Intervention  MAC (N = 16) | Control  TAU (N = 5) | P Value |
| --- | --- | --- | --- |
| Rapport (Child) |  |  | 0.1 |
| 0 | 11 (69%) | 1 (20%) |  |
| 1 | 0 (0%) | 0 (0%) |  |
| 2 | 2 (13%) | 2 (40%) |  |
| 3 | 3 (19%) | 2 (40%) |  |
| Rapport (Mother) |  |  | 1.0 |
| 0 | 14 (88%) | 5 (100%) |  |
| 1 | 1 (6%) | 0 (0%) |  |
| 2 | 1 (6%) | 0 (0%) |  |
| Rapport (Parents) |  |  |  |
| 0 | 15 (94%) | 5 (100%) | 1.0 |
| 1 | 1 (6%) | 0 (0%) |  |
| Rapport (Father) |  |  |  |
| 0 | 16 (100%) | 4 (80%) | 0.5 |
| 1 | 0 (6%) | 1 (20%) |  |
| Rapport (School) |  |  |  |
| 0 | 16 (100%) | 5 (100%) |  |
| Rapport (Family) |  |  |  |
| 0 | 16(100%) | 5 (100%) |  |
| Rapport (Other) |  |  |  |
| 0 | 16 (100%) | 5 (100%) |  |
| Risk (Child) |  |  | 0.5 |
| 0 | 7 (44%) | 1 (20%) |  |
| 1 | 4 (25%) | 2 (40%) |  |
| 2 | 2 (13%) | 0 (0%) |  |
| 3 | 3 (19%) | 2 (40%) |  |
| Risk (Mother) |  |  | 1.0 |
| 0 | 12 (75%) | 4 (80%) |  |
| 1 | 1 (6%) | 0 (0%) |  |
| 2 | 2 (13%) | 0 (0%) |  |
| 3 | 1 (6%) | 1 (20%) |  |
| Risk (School) |  |  |  |
| 0 | 14 (88%) | 5 (100%) | 1.0 |
| 1 | 1 (6%) | 0 (0%) |  |
| 2 | 0 (0%) | 0 (0%) |  |
| 3 | 1 (6%) | 0 (0%) |  |
| Risk (Father) |  |  | 0.5 |
| 0 | 16 (100%) | 4 (80%) |  |
| 1 | 0 (0%) | 1 (20%) |  |
| Risk (Parents) |  |  |  |
| 0 | 16 (100%) | 5 (100%) |  |
| Risk (Family) |  |  |  |
| 0 | 16 (100%) | 5 (100%) |  |
| Risk (Other) |  |  | 1.0 |
| 0 | 15 (94%) | 5 (80%) |  |
| 1 | 0 (0%) | 0 (0%) |  |
| 2 | 0 (0%) | 0 (0%) |  |
| 3 | 1 (6%) | 1 (20%) |  |
| Functioning (Child) |  |  | 0.2 |
| 0 | 15 (94%) | 3 (60%) |  |
| 1 | 1 (6%) | 1 (20%) |  |
| 2 | 0 (0%) | 0 (0%) |  |
| 3 | 0 (0%) | 1 (20%) |  |
| Functioning (Mother) |  |  | 0.5 |
| 0 | 15 (96%) | 4 (80%) |  |
| 1 | 1 (6%) | 0 (0%) |  |
| 2 | 0 (0%) | 1 (20%) |  |
| Functioning (Father) |  |  | 0.5 |
| 0 | 16 (100%) | 4 (80%) |  |
| 1 | 0 (0%) | 1 (20%) |  |
| Functioning (Parents) |  |  |  |
| 0 | 16 (100%) | 5 (100%) |  |
| Functioning (Family) |  |  |  |
| 0 | 16 (100%) | 5 (100%) |  |
| Functioning (School) |  |  |  |
| 0 | 16 (100%) | 5 (100%) |  |
| Functioning (Other) |  |  |  |
| 0 | 16 (100%) | 5 (100%) |  |
| Mental Health (Child) |  |  | 0.03 |
| 0 | 13 (82%) | 1 (20%) |  |
| 1 | 1 (6%) | 2 (40%) |  |
| 2 | 2 (13%) | 1 (20%) |  |
| 3 | 0 (0%) | 1 (20%) |  |
| Mental Health (Mother) |  |  | 0.5 |
| 0 | 16 (100%) | 4 (80%) |  |
| 1 | 0 (0%) | 0 (0%) |  |
| 2 | 0 (0%) | 1 (20%) |  |
| Mental Health (Father) |  |  |  |
| 0 | 16 (100%) | 5 (100%) |  |
| Mental Health (Parents) |  |  |  |
| 0 | 16 (100%) | 5 (100%) |  |
| Mental Health (School) |  |  |  |
| 0 | 16 (100%) | 5 (100%) |  |
| Mental Health (Family) |  |  |  |
| 0 | 16 (100%) | 5 (100%) |  |
| Mental Health (Other) |  |  |  |
| 0 | 16 (100%) | 5 (100%) |  |
| Motivational Interviewing (Child) |  |  | 0.9 |
| 0 | 12 (75%) | 3 (60%) |  |
| 1 | 1 (6%) | 1 (20%) |  |
| 2 | 3 (19%) | 1 (20%) |  |
| Motivational Interviewing (Mother) |  |  | 1.0 |
| 0 | 15 (94%) | 5 (100%) |  |
| 1 | 1 (6%) | 0 (0%) |  |
| Motivational Interviewing (Father) |  |  |  |
| 0 | 16 (100%) | 5 (100%) |  |
| Motivational Interviewing (Parents) |  |  |  |
| 0 | 16 (100%) | 5 (100%) |  |
| Motivational Interviewing (School) |  |  |  |
| 0 | 16 (100%) | 5 (100%) |  |
| Motivational Interviewing (Family) |  |  |  |
| 0 | 16 (100%) | 5 (100%) |  |
| Motivational Interviewing (Other) |  |  |  |
| 0 | 16 (100%) | 5 (100%) |  |
| Supportive Listening (Child) |  |  |  |
| 0 | 5 (31%) | 2 (40%) |  |
| 1 | 1 (6%) | 0 (0%) |  |
| 2 | 5 (31%) | 1 (20%) |  |
| 3 | 5 (31%) | 2 (40%) |  |
| Supportive Listening (Mother) |  |  | 1.0 |
| 0 | 13 (81%) | 4 (20%) |  |
| 1 | 0 (0%) | 0 (0%) |  |
| 2 | 3 (19%) | 1 (80%) |  |
| Supportive Listening (Father) |  |  | 0.5 |
| 0 | 16 (100%) | 4 (80%) |  |
| 1 | 0 (0%) | 1 (20%) |  |
| Supportive Listening (Parents) |  |  |  |
| 0 | 16 (100%) | 5 (100%) |  |
| Supportive Listening (School) |  |  |  |
| 0 | 16 (100%) | 5 (100%) |  |
| Supportive Listening (School) |  |  |  |
| 0 | 16 (100%) | 5 (100%) |  |
| Emotional Processing (Child) |  |  | 0.1 |
| 0 | 11 (69%) | 1 (20%) |  |
| 1 | 1 (6%) | 1 (20%) |  |
| 2 | 3 (19%) | 2 (40%) |  |
| 3 | 1 (6%) | 1 (20%) |  |
| Emotional Processing (Mother) |  |  | 0.1 |
| 0 | 16 (100%) | 3 (60%) |  |
| 1 | 0 (0%) | 1 (20%) |  |
| 2 | 0 (0%) | 1 (20%) |  |
| Emotional Processing (Father) |  |  |  |
| 0 | 16 (100%) | 5 (100%) |  |
| Emotional Processing (Parents) |  |  |  |
| 0 | 16 (100%) | 5 (100%) |  |
| Emotional Processing (School) |  |  |  |
| 0 | 16 (100%) | 5 (100%) |  |
| Emotional Processing (Family) |  |  |  |
| 0 | 16 (100%) | 5 (100%) |  |
| Emotional Processing (Other) |  |  |  |
| 0 | 16 (100%) | 5 (100%) |  |
| Problem Solving (Child) |  |  | 0.7 |
| 0 | 7 (44%) | 2 (40%) |  |
| 1 | 2 (13%) | 2 (40%) |  |
| 2 | 5 (31%) | 1 (20%) |  |
| 3 | 2 (13%) | 0 (0%) |  |
| Problem-Solving (Mother) |  |  | 1.0 |
| 0 | 14 (88%) | 4 (80%) |  |
| 1 | 0 (0%) | 1 (20%) |  |
| 2 | 1 (6%) | 0 (0%) |  |
| 3 | 1 (6%) | 0 (0%) |  |
| Problem Solving (Father) |  |  |  |
| 0 | 16 (100%) | 5 (100%) |  |
| Problem Solving (Parents) |  |  | 0.5 |
| 0 | 16 (100%) | 4 (80%) |  |
| 1 | 0 (0%) | 1 (20%) |  |
| Problem Solving (School) |  |  | 0.5 |
| 0 | 16 (100%) | 4 (80%) |  |
| 1 | 0 (0%) | 1 (20%) |  |
| Problem-Solving (Family) |  |  |  |
| 0 | 16 (100%) | 5 (100%) |  |
| Problem-Solving (Other) |  |  |  |
| 0 | 16 (100%) | 5 (100%) |  |
| Psycho-Education (Child) |  |  | 0.5 |
| 0 | 10 (63%) | 2 (40%) |  |
| 1 | 1 (6%) | 1 (20%) |  |
| 2 | 5 (31%) | 1 (20%) |  |
| 3 | 0 (0%) | 1 (20%) |  |
| Psycho-Education (Mother) |  |  | 1.0 |
| 0 | 13 (81%) | 4 (80%) |  |
| 1 | 0 (0%) | 0 (0%) |  |
| 2 | 3 (19%) | 1 (20%) |  |
| Psycho-Education (Mother) |  |  | 1.0 |
| 0 | 13 (81%) | 4 (80%) |  |
| 1 | 0 (0%) | 0 (0%) |  |
| 2 | 3 (19%) | 1 (20%) |  |
| Psycho-Education (Parents) |  |  | 0.9 |
| 0 | 15 (94%) | 4 (80%) |  |
| 1 | 1 (6%) | 1 (20%) |  |
| Psycho-Education (Father) |  |  | 0.5 |
| 0 | 16 (100%) | 4 (80%) |  |
| 1 | 0 (0%) | 1 (20%) |  |
| Psycho-Education (Family) |  |  |  |
| 0 | 16 (100%) | 5 (100%) |  |

| Psycho-Education (Other) |  | | |  |  |
| --- | --- | --- | --- | --- | --- |
| 0 | 16 (100%) | | | 5 (100%) |  |
| Goal Setting (Child) |  | | |  | 0.6 |
| 0 | 10 (63%) | | | 2 (40%) |  |
| 1 | 2 (13%) | | | 1 (20%) |  |
| 2 | 3 (19%) | | | 2 (40%) |  |
| 3 | 1 (6%) | | | 0 (0%) |  |
| Goal Setting (Mother) |  | | |  | 1.0 |
| 0 | 14 (88%) | | | 4 (80%) |  |
| 1 | 2 (13%) | | | 1 (20%) |  |
| Goal Setting (School) |  | | |  | 0.9 |
| 0 | 15 (94%) | | | 4 (80%) |  |
| 1 | 1 (6%) | | | 1 (20%) |  |
| Goal Setting (Father) |  | | |  | 0.5 |
| 0 | 16 (100%) | | | 4 (80%) |  |
| 1 | 0 (0%) | | | 1 (20%) |  |
| Goal Setting (Parents) |  | | |  |  |
| 0 | 16 (100%) | | | 5 (100%) |  |
| Goal Setting (Family) |  | | |  |  |
| 0 | 16 (100%) | | | 5 (100%) |  |
| Goal Setting (Other) |  | | |  |  |
| 0 | 16 (100%) | | | 5 (100%) |  |
| Activity Scheduling (Child) |  | | |  | 0.6 |
| 0 | 10 (63%) | | | 2 (40%) |  |
| 1 | 1 (6%) | | | 1 (20%) |  |
| 2 | 2 (13%) | | | 1 (20%) |  |
| 3 | 3 (19%) | | | 1 (20%) |  |
| Activity Scheduling (Mother) |  | | |  | 0.9 |
| 0 | 15 (94%) | | | 4 (80%) |  |
| 1 | 0 (0%) | | | 0 (0%) |  |
| 2 | 1 (6%) | | | 1 (20%) |  |
| Activity Scheduling (Father) |  | | |  | 0.1 |
| 0 | 16 (100%) | | | 5 (100%) |  |
| Activity Scheduling (Parents) |  | | |  |  |
| 0 | 16 (100%) | | | 5 (100%) |  |
| Activity Scheduling (School) |  | | |  |  |
| 0 | 16 (100%) | | | 5 (100%) |  |
| Activity Scheduling (Family) |  | | |  | 0.5 |
| 0 | 16 (100%) | | | 4 (80%) |  |
| 1 | 0 (0%) | | | 1 (20%) |  |
| Activity Scheduling (Other) |  | | |  | 1.0 |
| 0 | 16 (100%) | | | 5 (100%) |  |
| Crisis Management (Child) |  | | |  | 0.1 |
| 0 | 14 (88%) | | | 4 (80%) |  |
| 1 | 2 (13%) | | | 0 (0%) |  |
| 2 | 0 (0%) | | | 1 (20%) |  |
| Crisis Management (Mother) |  | | |  | 0.6 |
| 0 | 15 (94%) | | | 5 (100%) |  |
| 1 | 0 (0%) | | | 0 (0%) |  |
| 2 | 1 (6%) | | | 0 (0%) |  |
| Crisis Management (Parents) |  | | |  | 1.0 |
| 0 | 15 (94%) | | | 5 (100%) |  |
| 1 | 0 (0%) | | | 0 (0%) |  |
| 2 | 1 (6%) | | | 0 (0%) |  |
| Crisis Management (Father) |  | | |  | 0.5 |
| 0 | 16 (100%) | | | 4 (80%) |  |
| 1 | 0 (0%) | | | 0 (0%) |  |
| 2 | 0 (0%) | | | 1 (20%) |  |
| Crisis Management (School) |  | | |  | 1.0 |
| 0 | 15 (94%) | | | 5 (100%) |  |
| 1 | 1 (6%) | | | 0 (0%) |  |
| Crisis Management (Family) |  | | |  |  |
| 0 | 16 (100%) | | | 5 (100%) |  |
| Crisis Management (Other) |  | | |  | 1.0 |
| 0 | 15 (94%) | | | 5 (100%) |  |
| 1 | 1 (6%) | | | 0 (0%) |  |
| Relapse Prevention (Child) |  | | |  | 1.0 |
| 0 | 9 (56%) | | | 3 (60%) |  |
| 1 | 1 (6%) | | | 0 (0%) |  |
| 2 | 4 (25%) | | | 1 (20%) |  |
| 3 | 2 (13%) | | | 1 (20%) |  |
| Relapse Prevention (Mother) |  | | |  | 0.8 |
| 0 | 31 (81%) | | | 5 (100%) |  |
| 1 | 2 (13%) | | | 0 (0%) |  |
| 2 | 1 (6%) | | | 0 (0%) |  |
| 3 |  | | |  |  |
| Relapse Prevention (Father) |  | | |  |  |
| 0 | 16 (100%) | | | 5 (100%) |  |
| Relapse Prevention (School) |  | | |  |  |
| 0 | 16 (100%) | | | 5 (100%) |  |
| Relapse Prevention (Parents) |  | | |  |  |
| 0 | 16 (100%) | | | 5 (100%) |  |
| Relapse Prevention (Family) |  | | |  |  |
| 0 | 16 (100%) | | | 5 (100%) |  |
| Relapse Prevention (Other) |  | | |  |  |
| 0 | 16 (100%) | | | 5 (100%) |  |
| Practical Support (Child) |  | | |  | 0.1 |
| 0 | 13 (81%) | | | 2 (40%) |  |
| 1 | 3 (19%) | | | 1 (20%) |  |
| 2 | 0 (0%) | | | 2 (40%) |  |
| Practical Support (Mother) |  | | |  | 0.9 |
| 0 | 15 (96%) | | | 4 (80%) |  |
| 1 | 1 (6%) | | | 1 (20%) |  |
| Practical Support (Father) |  | | |  |  |
| 0 | 16 (100%) | | | 5 (100%) |  |
| Practical Support (Parents) |  | | |  |  |
| 0 | 16 (100%) | | | 5 (100%) |  |
| Practical Support (School) |  | | |  |  |
| 0 | 16 (100%) | | | 5 (100%) |  |
| Practical Support (Family) |  | | |  |  |
| 0 | 16 (100%) | | | 5 (100%) |  |
| Practical Support (Other) |  | | |  |  |
| 0 | 16 (100%) | | | 5 (100%) |  |
| Supervision (Child) |  | | |  | 0.5 |
| 0 | 16 (100%) | | | 4 (80%) |  |
| 1 | 0 (0%) | | | 1 (20%) |  |
| Supervision (Mother) |  | | |  |  |
| 0 | 16 (100%) | | | 5 (100%) |  |
| Supervision (Father) |  | | |  |  |
| 0 | 16 (100%) | | | 5 (100%) |  |
| Supervision (Parents) |  | | |  |  |
| 0 | 16 (100%) | | | 5 (100%) |  |
| Supervision (School) |  | | |  |  |
| 0 | 16 (100%) | | | 5 (100%) |  |
| Supervision (Family) |  | | |  |  |
| 0 | 16 (100%) | | | 5 (100%) |  |
| Supervision (Other) |  | | |  |  |
| 0 | 16 (100%) | | | 5 (100%) |  |
| Monitoring Training (Child) |  | | |  | 0.5 |
| 0 | 15 (94%) | | | 4 (80%) |  |
| 1 | 1 (6%) | | | 1 (20%) |  |
| Monitoring Training (Mother) |  | | |  |  |
| 0 | 15 (94%) | | | 5 (100%) | 1.0 |
| 1 | 1 (6%) | | | 0 (0%) |  |
| Monitoring Training (Father) |  | | |  |  |
| 0 | 16 (100%) | | | 5 (100%) |  |
| Monitoring Training (Parents) |  | | |  |  |
| 0 | 15 (94%) | | | 5 (100%) | 1.0 |
|  | 1 (6%) | | | 0 (0%) |  |
| Monitoring Training (School) |  | | |  |  |
| 0 | 16 (100%) | | | 5 (100%) |  |
| Monitoring Training (Family) |  | | |  |  |
| 0 | 16 (100%) | | | 5 (100%) |  |
| Monitoring Training (Other) |  | | |  |  |
| 0 | 16 (100%) | | | 5 (100%) |  |
| Interpretation (Child) | |  |  | | 0.1 |
| 0 | | 16 (100%) | 3 (60%) | |  |
| 1 | | 0 (0%) | 1 (20%) | |  |
| 2 | | 0 (0%) | 1 (20%) | |  |
| Interpretation (Mother) | |  |  | | 0.5 |
| 0 | | 16 (100%) | 4 (80%) | |  |
| 1 | | 0 (0%) | 1 (20%) | |  |
| Interpretation (Father) | |  |  | | 0.5 |
| 0 | | 16 (100%) | 4 (80%) | |  |
| 1 | | 0 (0%) | 1 (20%) | |  |
| Interpretation (Parents) | |  |  | |  |
| 0 | | 16 (100%) | 5 (100%) | |  |
| Interpretation (School) | |  |  | |  |
| 0 | | 16 (100%) | 5 (100%) | |  |
| Interpretation (Family) | |  |  | |  |
| 0 | | 16(100%) | 5 (100%) | |  |
| Interpretation (Other) | |  |  | |  |
| 0 | | 16(100%) | 5 (100%) | |  |
| Catharsis (Child) | |  |  | | 0.5 |
| 0 | | 16 (100%) | 4 (80%) | |  |
| 1 | | 0 (0%) | 1 (20%) | |  |
| Catharsis (Mother) | |  |  | |  |
| 0 | | 16 (100%) | 5 (100%) | |  |
| Catharsis (Father) | |  |  | |  |
| 0 | | 16 (100%) | 5 (100%) | |  |
| Catharsis (Parents) | |  |  | |  |
| 0 | | 16 (100%) | 5 (100%) | |  |
| Catharsis (School) | |  |  | |  |
| 0 | | 16 (100%) | 5 (100%) | |  |
| Catharsis (Family) | |  |  | |  |
| 0 | | 16 (100%) | 5 (100%) | |  |
| Catharsis (Other) | |  |  | |  |
| 0 | | 16 (100%) | 5 (100%) | |  |
| Insight Building (Child) | |  |  | | 0.9 |
| 0 | | 11 (69%) | 3 (60%) | |  |
| 1 | | 1 (6%) | 1 (20%) | |  |
| 2 | | 4 (25%) | 0 (0%) | |  |
| 3 | | 0 (0%) | 1 (20%) | |  |
| Insight Building (Mother) | |  |  | | 0.9 |
| 0 | | 15 (94%) | 4 (80%) | |  |
| 1 | | 0 (0%) | 0 (0%) | |  |
| 2 | | 1 (6%) | 1 (20%) | |  |
| Insight Building (Father) | |  |  | |  |
| 0 | | 16 (100%) | 5 (100%) | |  |
| Insight Building (Parents) | |  |  | |  |
| 0 | | 16 (100%) | 5 (100%) | |  |
| Insight Building (School) | |  |  | |  |
| 0 | | 16 (100%) | 5 (100%) | |  |
| Insight Building (Family) | |  |  | | 0.5 |
| 0 | | 16 (100%) | 4 (80%) | |  |
| 1 | | 0 (0%) | 1 (20%) | |  |
| Insight Building (Other) | |  |  | |  |
| 0 | | 16 (100%) | 5 (100%) | |  |
| Mindfulness (Child) | |  |  | | 0.3 |
| 0 | | 12 (75%) | 2 (40%) | |  |
| 1 | | 2 (13%) | 2 (40%) | |  |
| 2 | | 2 (13%) | 1 (20%) | |  |
| Mindfulness (Mother) | |  |  | |  |
| 0 | | 16 (100%) | 5 (100%) | |  |
| Mindfulness (Father) | |  |  | |  |
| 0 | | 16 (100%) | 5 (100%) | |  |
| Mindfulness (Parents) | |  |  | |  |
| 0 | | 16 (100%) | 5 (100%) | |  |
| Mindfulness (School) | |  |  | |  |
| 0 | | 16 (100%) | 5 (100%) | |  |
| Mindfulness (Family) | |  |  | |  |
| 0 | | 16 (100%) | 5 (100%) | |  |
| Mindfulness (Other) | |  |  | |  |
| 0 | | 16 (100%) | 5 (100%) | |  |
| Logical Consequences (Child) | |  |  | | 1.0 |
| 0 | | 14 (88%) | 4 (80%) | |  |
| 1 | | 2 (13%) | 1 (20%) | |  |
| Logical Consequences (Mother) | |  |  | | 1.0 |
| 0 | | 15 (94%) | 5 (100%) | |  |
| 1 | | 1 (6%) | 0 (0%) | |  |
| Logical Consequences (Father) | |  |  | |  |
| 0 | | 16 (100%) | 5 (100%) | |  |
| Logical Consequences (Parents) | |  |  | |  |
| 0 | | 16 (100%) | 5 (100%) | |  |
| Logical Consequences (School) | |  |  | |  |
| 0 | | 16 (100%) | 5 (100%) | |  |
| Logical Consequences (Family) | |  |  | |  |
| 0 | | 16 (100%) | 5 (100%) | |  |
| Logical Consequences (Other) | |  |  | |  |
| 0 | | 16 (100%) | 5 (100%) | |  |
| Praise (Child) | |  |  | | 1.0 |
| 0 | | 9 (56%) | 3 (60%) | |  |
| 1 | | 2 (13%) | 0 (0%) | |  |
| 2 | | 2 (13%) | 1 (20%) | |  |
| 3 | | 3 (19%) | 1 (20%) | |  |
| Praise (Mother) | |  |  | | 0.9 |
| 0 | | 15 (94%) | 4 (80%) | |  |
| 1 | | 0 (0%) | 1 (20%) | |  |
| 2 | | 1 (6%) | 0 (0%) | |  |
| Praise (Father) | |  |  | |  |
| 0 | | 16 (100%) | 5 (100%) | |  |
| Praise (Parents) | |  |  | |  |
| 0 | | 16 (100%) | 5 (100%) | |  |
| Praise (School) | |  |  | |  |
| 0 | | 16 (100%) | 5 (100%) | |  |
| Praise (Family) | |  |  | |  |
| 0 | | 16 (100%) | 5 (100%) | |  |
| Praise (Other) | |  |  | |  |
| 0 | | 16 (100%) | 5 (100%) | |  |
| Modelling (Child) | |  |  | | 0.2 |
| 0 | | 15 (100%) | 3 (60%) | |  |
| 1 | | 0 (0%) | 1 (20%) | |  |
| 2 | | 1 (6%) | 0 (0%) | |  |
| 3 | | 0 (0%) | 1 (20%) | |  |
| Modelling (Mother) | |  |  | | 0.9 |
| 0 | | 15 (94%) | 4 (80%) | |  |
| 1 | | 1 (6%) | 1 (20%) | |  |
| Modelling (Father) | |  |  | | 0.5 |
| 0 | | 16 (100%) | 4 (80%) | |  |
| 1 | | 0 (0%) | 1 (20%) | |  |
| Modelling (Parents) | |  |  | | 1.0 |
| 0 | | 16 (100%) | 5 (100%) | |  |
| Modelling (School) | |  |  | |  |
| 0 | | 16 (100%) | 5 (100%) | |  |
| Modelling (Family) | |  |  | |  |
| 0 | | 16 (100%) | 5 (100%) | |  |
| Modelling (Other) | |  |  | |  |
| 0 | | 16 (100%) | 5 (100%) | |  |
| Self-Reward Training (Child) | |  |  | |  |
| 0 | | 13 (81%) | 4 (80%) | |  |
| 1 | | 1 (6%) | 1 (20%) | |  |
| 2 | | 1 (6%) | 0 (0%) | |  |
| 3 | | 1 (6%) | 0 (0%) | |  |
| P=1.0 | |  |  | |  |
| Self-Reward Training (Mother) | |  |  | | 1.0 |
| 0 | | 15 (94%) | 5 (100%) | |  |
| 1 | | 0 (0%) | 0 (0%) | |  |
| 2 | | 1 (6%) | 0 (0%) | |  |
| Self-Reward Training (Father) | |  |  | |  |
| 0 | | 16 (100%) | 5 (100%) | |  |
| Self-Reward Training (Parents) | |  |  | |  |
| 0 | | 16 (100%) | 5 (100%) | |  |
| Self-Reward Training (School) | |  |  | |  |
| 0 | | 16 (100%) | 5 (100%) | |  |
| Self-Reward Training (Family) | |  |  | |  |
| 0 | | 16 (100%) | 5 (100%) | |  |
| Self-Reward Training (Other) | |  |  | |  |
| 0 | | 16 (100%) | 5 (100%) | |  |
| Stimulus Control (Child) | |  |  | | 0.1 |
| 0 | | 16 (100%) | 3 (60%) | |  |
| 1 | | 0 (0%) | 1 (20%) | |  |
| 2 | | 0 (0%) | 0 (0%) | |  |
| 3 | | 0 (0%) | 1 (20%) | |  |
| Stimulus Control (Mother) | |  |  | | 0.5 |
| 0 | | 16 (100%) | 4 (80%) | |  |
| 1 | | 0 (0%) | 1 (20%) | |  |
| Stimulus Control (Father) | |  |  | | 0.5 |
| 0 | | 16 (100%) | 4 (80%) | |  |
| 1 | | 0 (0%) | 1 (20%) | |  |
| Stimulus Control (Father) | |  |  | | 0.5 |
| 0 | | 16 (100%) | 4 (80%) | |  |
| 1 | | 0 (0%) | 1 (20%) | |  |
| Stimulus Control (Parents) | |  |  | |  |
| 0 | | 16 (100%) | 5 (100%) | |  |
| Stimulus Control (School) | |  |  | |  |
| 0 | | 16 (100%) | 5 (100%) | |  |

| Stimulus Control (Family) |  |  | |  | |
| --- | --- | --- | --- | --- | --- |
| 0 | 16(100%) | 5 (100%) | |  | |
| Stimulus Control (Other) |  |  | |  | |
| 0 | 16(100%) | 5 (100%) | |  | |
| Time-Out (Child) |  |  | | 0.9 | |
| 0 | 15 (94%) | 4 (80%) | |  | |
| 1 | 1 (6%) | 1 (20%) | |  | |
| Time-Out (Mother) |  |  | |  | |
| 0 | 16 (100%) | 5 (100%) | |  | |
| Time-Out (Father) |  |  | |  | |
| 0 | 16 (100%) | 5 (100%) | |  | |
| Time-Out (Parents) |  |  | |  | |
| 0 | 16 (100%) | 5 (100%) | |  | |
| Time-Out (School) |  |  | |  | |
| 0 | 16 (100%) | 5 (100%) | |  | |
| Time-Out (Family) |  |  | |  | |
| 0 | 16 (100%) | 5 (100%) | |  | |
| Time-Out (Other) |  |  | |  | |
| 0 | 16 (100%) | 5 (100%) | |  | |
| Response Cost (Child) |  |  | | 0.5 | |
| 0 | 16 (100%) | 4 (80%) | |  | |
| 1 | 0 (0%) | 1 (20%) | |  | |
| Response Cost (Mother) |  |  | |  | |
| 0 | 16 (100%) | 5 (100%) | |  | |
| Response Cost (Father) |  |  | |  | |
| 0 | 16 (100%) | 5 (100%) | |  | |
| Response Cost (Parents) |  |  | |  | |
| 0 | 16 (100%) | 5 (100%) | |  | |
| Response Cost (School) |  |  | |  | |
| 0 | 16 (100%) | 5 (100%) | |  | |
| Response Cost (Family) |  |  | | 0.5 | |
| 0 | 16 (100%) | 4 (80%) | |  | |
| 1 | 0 (0%) | 1 (20%) | |  | |
| Response Cost (Other) |  |  | |  | |
| 0 | 16 (100%) | 5 (100%) | |  | |
| Response Prevention (Child) |  |  | | 0.3 | |
| 0 | 15 (94%) | 3 (60%) | |  | |
| 1 | 0 (0%) | 1 (20%) | |  | |
| 2 | 1 (6%) | 1 (20%) | |  | |
| Response Prevention (Mother) |  |  | | 0.5 | |
| 0 | 16 (100%) | 4 (80%) | |  | |
| 1 | 0 (0%) | 1 (20%) | |  | |
| Response Prevention (Father) |  |  | | 0.5 | |
| 0 | 16 (100%) | 4 (80%) | |  | |
| 1 | 0 (0%) | 1 (20%) | |  | |
| Response Prevention (Parents) |  |  | |  | |
| 0 | 16 (100%) | 5 (100%) | |  | |
| Response Prevention (School) |  |  | |  | |
| 0 | 16 (100%) | 5 (100%) | |  | |
| Response Prevention (Family) |  |  | |  | |
| 0 | 16 (100%) | 5 (100%) | |  | |
| Response Prevention (Other) |  |  | |  | |
| 0 | 16 (100%) | 5 (100%) | |  | |
| Tangential Rewards (Child) |  |  | | 1.0 | |
| 0 | 14 (88%) | 4 (80%) | |  | |
| 1 | 1 (6%) | 1 (20%) | |  | |
| 2 | 1 (6%) | 0 (0%) | |  | |
| Tangential Rewards (Mother) |  |  | | 0.9 | |
| 0 | 15 (96%) | 4 (80%) | |  | |
| 1 | 0 (0%) | 0 (0%) | |  | |
| 2 | 1 (6%) | 1 (20%) | |  | |
| Tangential Rewards (Father) |  |  | |  | |
| 0 | 16 (100%) | 5 (100%) | |  | |
| Tangential Rewards (Parents) |  |  | |  | |
| 0 | 16 (100%) | 5 (100%) | |  | |
| Tangential Rewards (School) |  |  | |  | |
| 0 | 16 (100%) | 5 (100%) | |  | |
| Tangential Rewards (Family) |  |  | | 0.5 | |
| 0 | 16 (100%) | 4 (80%) | |  | |
| 1 | 0 (0%) | 0 (0%) | |  | |
| 2 | 0 (0%) | 1 (20%) | |  | |
| Tangential Rewards (Other) |  |  | |  | |
| 0 | 16 (100%) | 5 (100%) | |  | |
| Relaxation (Child) |  |  | | 0.9 | |
| 0 | 12 (75%) | 3 (60%) | |  | |
| 1 | 0 (0%) | 1 (20%) | |  | |
| 2 | 4 (25%) | 1 (20%) | |  | |
| Relaxation (Mother) |  |  | | 0.9 | |
| 0 | 15 (94%) | 5 (100%) | |  | |
| 1 | 0 (0%) | 0 (0%) | |  | |
| 2 | 1 (6%) | 0 (0%) | |  | |
| Relaxation (Father) |  |  | |  | |
| 0 | 16 (100%) | 5 (100%) | |  | |
| Relaxation (Parents) |  |  | |  | |
| 0 | 16 (100%) | 5 (100%) | |  | |
| Relaxation (School) |  |  | |  | |
| 0 | 16 (100%) | 5 (100%) | |  | |
| Relaxation (Family) |  |  | |  | |
| 0 | 16 (100%) | 5 (100%) | |  | |
| Relaxation (Other) |  |  | |  | |
| 0 | 16 (100%) | 5 (100%) | |  | |
| Guided Imagery (Child) |  |  | | 0.9 | |
| 0 | 15 (80%) | 4 (80%) | |  | |
| 1 | 1 (20%) | 1 (20%) | |  | |
| Guided Imagery (Mother) |  |  | |  | |
| 0 | 16 (100%) | 5 (100%) | |  | |
| Guided Imagery (Father) |  |  | |  | |
| 0 | 16 (100%) | 5 (100%) | |  | |
| Guided Imagery (Parents) |  |  | |  | |
| 0 | 16 (100%) | 5 (100%) | |  | |
| Guided Imagery (School) |  |  | |  | |
| 0 | 16 (100%) | 5 (100%) | |  | |
| Guided Imagery (Family) |  |  | |  | |
| 0 | 16 (100%) | 5 (100%) | |  | |
| Guided Imagery (Other) |  |  | |  | |
| 0 | 16 (100%) | 5 (100%) | |  | |
| Cognitive/Coping (Child) |  |  | | 0.1 | |
| 0 | 11 (69%) | 1 (20%) | |  | |
| 1 | 3 (19%) | 2 (40%) | |  | |
| 2 | 0 (0%) | 1 (20%) | |  | |
| 3 | 2 (6%) | 1 (20%) | |  | |
| Cognitive/Coping (Mother) |  |  | | 1.0 | |
| 0 | 15 (94%) | 5 (100%) | |  | |
| 1 | 0 (0%) | 0 (0%) | |  | |
| 2 | 0 (0%) | 0 (0%) | |  | |
| 3 | 1 (6%) | 0 (0%) | |  | |
| Cognitive/Coping (Father) |  |  | | 0.5 | |
| 0 | 16 (100%) | 4 (80%) | |  | |
| 1 | 0 (0%) | 0 (0%) | |  | |
| 2 | 0 (0%) | 1 (20%) | |  | |
| Cognitive/Coping (Parents) |  |  | |  | |
| 0 | 16 (100%) | 5 (100%) | |  | |
| Cognitive/Coping (School) |  |  | |  | |
| 0 | 16 (100%) | 5 (100%) | |  | |
| Cognitive/Coping (Family) |  |  | |  | |
| 0 | 16 (100%) | 5 (100%) | |  | |
| Cognitive/Coping (Other) |  |  | |  | |
| 0 | 16 (100%) | 5 (100%) | |  | |
| Exposure (Child) |  | |  | | 0.5 |
| 0 | 14 (88%) | | 3 (60%) | |  |
| 1 | 1 (6%) | | 2 (40%) | |  |
| 2 | 1 (6%) | | 0 (0%) | |  |
| Exposure (Mother) |  | |  | | 1.0 |
| 0 | 15 (94%) | | 5 (100%) | |  |
| 1 | 0 (0%) | | 0 (0%) | |  |
| 2 | 1 (6%) | | 0 (0%) | |  |
| Exposure (Father) |  | |  | |  |
| 0 | 16 (100%) | | 5 (100%) | |  |
| Exposure (Parents) |  | |  | |  |
| 0 | 16 (100%) | | 5 (100%) | |  |
| Exposure (School) |  | |  | |  |
| 0 | 16 (100%) | | 5 (100%) | |  |
| Exposure (Family) |  | |  | |  |
| 0 | 16(100%) | | 5 (100%) | |  |
| Exposure (Other) |  | |  | |  |
| 0 | 16(100%) | | 5 (100%) | |  |
| Communication Skills (Child) |  | |  | | 0.6 |
| 0 | 11 (69%) | | 3 (60%) | |  |
| 1 | 5 (31%) | | 1 (20%) | |  |
| 2 | 0 (0%) | | 1 (20%) | |  |
| Communication Skills (Mother) |  | |  | | 1.0 |
| 0 | 15 (94%) | | 5 (100%) | |  |
| 1 | 1 (6%) | | 0 (0%) | |  |
| Communication Skills (Father) |  | |  | |  |
| 0 | 16 (100%) | | 5 (100%) | |  |
| Communication Skills (Parents) |  | |  | |  |
| 0 | 16 (100%) | | 5 (100%) | |  |
| Communication Skills (School) |  | |  | |  |
| 0 | 16 (100%) | | 5 (100%) | |  |
| Communication Skills (Family) |  | |  | |  |
| 0 | 15 (94%) | | 5 (100%) | | 1.0 |
| 1 | 1 (6%) | | 0 (0%) | |  |
| Communication Skills (Other) |  | |  | |  |
| 0 | 16 (100%) | | 5 (100%) | |  |
| Social Skills (Child) |  | |  | | 0.9 |
| 0 | 15 (94%) | | 4 (80%) | |  |
| 1 | 1 (6%) | | 1 (20%) | |  |
| Social Skills (Mother) |  | |  | |  |
| 0 | 16 (100%) | | 5 (100%) | |  |
| Social Skills (Father) |  | |  | |  |
| 0 | 16 (100%) | | 5 (100%) | |  |
| Social Skills (Parents) |  | |  | |  |
| 0 | 16 (100%) | | 5 (100%) | |  |
| Social Skills (School) |  | |  | |  |
| 0 | 16 (100%) | | 5 (100%) | |  |
| Social Skills (Family) |  | |  | |  |
| 0 | 16 (100%) | | 5 (100%) | |  |
| Social Skills (Other) |  | |  | |  |
| 0 | 16 (100%) | | 5 (100%) | |  |
| Assertiveness (Child) |  | |  | | 1.0 |
| 0 | 14 (88%) | | 3 (60%) | |  |
| 1 | 2 (13%) | | 1 (20%) | |  |
| Assertiveness (Mother) |  | |  | |  |
| 0 | 15 (94%) | | 5 (100%) | | 1.0 |
| 1 | 1 (6%) | | 0 (0%) | |  |
| Assertiveness (Father) |  | |  | |  |
| 0 | 16 (100%) | | 5 (100%) | |  |
| Assertiveness (Parents) |  | |  | |  |
| 0 | 16 (100%) | | 5 (100%) | |  |
| Assertiveness (School) |  | |  | |  |
| 0 | 16 (100%) | | 5 (100%) | |  |
| Assertiveness (Family) |  | |  | |  |
| 0 | 16 (100%) | | 5 (100%) | |  |
| Assertiveness (Other) |  | |  | |  |
| 0 | 16 (100%) | | 5 (100%) | |  |
| Limit-Setting (Child) |  | |  | | 0.5 |
| 0 | 16 (100%) | | 4 (80%) | |  |
| 1 | 0 (0%) | | 1 (20%) | |  |
| Limit-Setting (Mother) |  | |  | |  |
| 0 | 16 (100%) | | 5 (100%) | |  |
| Limit Setting (Father) |  | |  | |  |
| 0 | 16 (100%) | | 5 (100%) | |  |
| Limit-Setting (Parents) |  | |  | |  |
| 0 | 16 (100%) | | 5 (100%) | |  |
| Limit-Setting (School) |  | |  | |  |
| 0 | 16 (100%) | | 5 (100%) | |  |
| Limit-Setting (Family) |  | |  | | 1.0 |
| 0 | 16 (100%) | | 4 (80%) | |  |
| 1 | 0 (0%) | | 0 (0%) | |  |
| 2 | 0 (0%) | | 1 (20%) | |  |
| Limit-Setting (Other) |  | |  | |  |
| 0 | 16 (100%) | | 5 (100%) | |  |
| Family Engagement (Child) |  | |  | | 0.1 |
| 0 | 13 (81%) | | 2 (40%) | |  |
| 1 | 2 (13%) | | 1 (20%) | |  |
| 2 | 1 (6%) | | 1 (20%) | |  |
| 3 | 0 (0%) | | 1 (20%) | |  |
| Family Engagement (Mother) |  | |  | | 0.1 |
| 0 | 15 (94%) | | 3 (60%) | |  |
| 1 | 1 (6%) | | 0 (0%) | |  |
| 2 | 0 (0%) | | 1 (20%) | |  |
| 3 | 0 (0%) | | 1 (20%) | |  |
| Family Engagement (Father) |  | |  | |  |
| 0 | 16 (100%) | | 5 (100%) | |  |
| Family Engagement (Parents) |  | |  | |  |
| 0 | 16 (100%) | | 5 (100%) | |  |
| Family Engagement (School) |  | |  | |  |
| 0 | 16 (100%) | | 5 (100%) | |  |
| Family Engagement (Family) |  | |  | |  |
| 0 | 15 (100%) | | 5 (100%) | | 1.0 |
| 1 | 1 (6%) | | 0 (0%) | |  |
| Family Engagement (Other) |  | |  | |  |
| 0 | 16 (100%) | | 5 (100%) | |  |
| Family Therapy (Child) |  | |  | | 0.1 |
| 0 | 16 (100%) | | 3 (60%) | |  |
| 1 | 0 (0%) | | 1 (20%) | |  |
| 2 | 0 (0%) | | 0 (0%) | |  |
| 3 | 0 (0%) | | 1 (20%) | |  |
| Family Therapy (Mother) |  | |  | | 0.5 |
| 0 | 16 (100%) | | 4 (80%) | |  |
| 1 | 0 (0%) | | 0 (0%) | |  |
| 2 | 0 (0%) | | 0 (0%) | |  |
| 3 | 0 (0%) | | 1 (20%) | |  |
| Family Therapy (Father) |  | |  | | 0.5 |
| 0 | 15 (94%) | | 4 (80%) | |  |
| 1 | 0 (0%) | | 0 (0%) | |  |
| 2 | 1 (6%) | | 0 (0%) | |  |
| 3 | 0 (0%) | | 1 (20%) | |  |
| Family Therapy (Parents) |  | |  | |  |
| 0 | 16 (100%) | | 5 (100%) | |  |
| Family Therapy (School) |  | |  | |  |
| 0 | 16 (100%) | | 5 (100%) | |  |
| Family Therapy (Family) |  | |  | | 0.5 |
| 0 | 15 (94%) | | 5 (100%) | |  |
| 1 | 0 (0%) | | 0 (0%) | |  |
| 2 | 1 (6%) | | 0 (0%) | |  |
| Family Therapy (Other) |  | |  | |  |
| 0 | 16 (100%) | | 5 (100%) | |  |
| Couples Therapy (Child) |  | |  | | 0.5 |
| 0 | 16 (100%) | | 4 (80%) | |  |
| 1 | 0 (0%) | | 1 (20%) | |  |
| Couples Therapy (Mother) |  | |  | |  |
| 0 | 16 (100%) | | 4 (80%) | |  |
| 1 | 0 (0%) | | 0 (0%) | |  |
| 2 | 0 (0%) | | 1 (20%) | |  |
| Couples Therapy (Father) |  | |  | | 0.5 |
| 0 | 16 (100%) | | 4 (80%) | |  |
| 1 | 0 (0%) | | 0 (0%) | |  |
| 2 | 0 (0%) | | 1 (20%) | |  |
| Couples Therapy (Parents) |  | |  | |  |
| 0 | 16 (100%) | | 5 (100%) | |  |
| Couples Therapy (School) |  | |  | |  |
| 0 | 16 (100%) | | 5 (100%) | |  |
| Couples Therapy (Family) |  | |  | | 1.0 |
| 0 | 15 (94%) | | 5 (100%) | |  |
| 1 | 0 (0%) | | 0 (0%) | |  |
| 2 | 1 (6%) | | 0 (0%) | |  |
| Couples Therapy (Other) |  | |  | |  |
| 0 | 16 (100%) | | 5 (100%) | |  |
| Play Therapy (Child) |  | |  | | 0.5 |
| 0 | 16 (100%) | | 4 (60%) | |  |
| 1 | 0 (0%) | | 1 (20%) | |  |
| Play Therapy (Mother) |  | |  | |  |
| 0 | 16 (100%) | | 5 (100%) | |  |
| Play Therapy (Father) |  | |  | |  |
| 0 | 16 (100%) | | 5 (100%) | |  |
| Play Therapy (Parents) |  | |  | |  |
| 0 | 16 (100%) | | 5 (100%) | |  |
| Play Therapy (Parents) |  | |  | |  |
| 0 | 16 (100%) | | 5 (100%) | |  |
| Play Therapy (Parents) |  | |  | |  |
| 0 | 16 (100%) | | 5 (100%) | |  |

| Play Therapy (Parents) |  |  |  |
| --- | --- | --- | --- |
| 0 | 16 (100%) | 5 (100%) |  |
| Play Therapy (School) |  |  |  |
| 0 | 16 (100%) | 5 (100%) |  |
| Play Therapy (Family) |  |  |  |
| 0 | 16(100%) | 5 (100%) |  |
| Play Therapy (Other) |  |  |  |
| 0 | 16(100%) | 5 (100%) |  |
| Therapeutic Letters (Child) |  |  | 0.6 |
| 0 | 16 (100%) | 4 (60%) |  |
| 1 | 0 (0%) | 1 (20%) |  |
| Therapeutic Letters (Mother) |  |  |  |
| 0 | 16 (100%) | 5 (100%) |  |
| Therapeutic Letters (Father) |  |  |  |
| 0 | 16 (100%) | 5 (100%) |  |
| Therapeutic Letters (Parents) |  |  |  |
| 0 | 16 (100%) | 5 (100%) |  |
| Therapeutic Letters (School) |  |  |  |
| 0 | 16 (100%) | 5 (100%) |  |
| Therapeutic Letters (Family) |  |  |  |
| 0 | 16 (100%) | 5 (100%) |  |
| Therapeutic Letters (Other) |  |  |  |
| 0 | 16 (100%) | 5 (100%) |  |
