## Appendix B TRS for "Mental health pathways and treatment as usual for young people experiencing depression in the United Kingdom: A mixed methods study"

CAMHS

Treatment Recording Sheet (TRS)

This tool is designed to record treatment provided by CAMHS for individual adolescents. Would you please record what has been conducted for this client SINCE YOU REFERRED THEM TO THE ATTEND STUDY. Please also record on the other side any critical incidents that have occurred. Notes on completion are provided at the back of the form.

| Client Initials |  | |
| --- | --- | --- |
| Number of Sessions |  | Clinician Name |

Primary Diagnosis: ______________________________________________________

Do you think the client also meets the diagnostic criteria for a depressive disorder?

☐ Yes	☐ No	☐ Unsure

### Week of Treatment

☐ 0	☐ 4	☐ 8	☐ 12	☐ 16	☐ 20	☐ 24	Date completed:___________________

### Clinical Discipline

☐ Psychological	☐ SW	☐ RPN	☐ OT	☐ Speech therapy

☐ Registrar	☐ Other

### Location (tick all that apply)

☐ Office	☐ Interview Room	☐ Home	☐ School

☐ Other________________________________________________

### Stage of Treatment:

| intake |  |  |  |  |  |  |  |  |  | closure |
| --- | --- | --- | --- | --- | --- | --- | --- | --- | --- | --- |

### Interventions tried (Definitions supplied at the back of this form)

|  | Key: Frequency  xxx = Frequent use; xx = Moderate use; x = Little use | Key: With Whom  C = Client; M = Mother; F = Father; P = Parents; Fam = Family; S = School; O = Other | | | | | |
| --- | --- | --- | --- | --- | --- | --- | --- |
|  | Interventions | Freq | With | Freq | With | Freq | With |
| 1 | Rapport building |  |  |  |  |  |  |
|  | Risk assessment |  |  |  |  |  |  |
|  | Functional analysis |  |  |  |  |  |  |
|  | Mental status exam |  |  |  |  |  |  |
| 2 | Motivational interviewing |  |  |  |  |  |  |
|  | Supportive listening |  |  |  |  |  |  |
|  | Emotional processing |  |  |  |  |  |  |
|  | Problem solving |  |  |  |  |  |  |
|  | Psychoeducation |  |  |  |  |  |  |
|  | Goal setting |  |  |  |  |  |  |
|  | Activity scheduling |  |  |  |  |  |  |
| 3 | Crisis management |  |  |  |  |  |  |
|  | Maintenance/relapse prevent |  |  |  |  |  |  |
|  | Practical support |  |  |  |  |  |  |
|  | Line of sight supervision |  |  |  |  |  |  |
|  | Monitoring training |  |  |  |  |  |  |
| 4 | Interpretation |  |  |  |  |  |  |
|  | Catharsis |  |  |  |  |  |  |
|  | Insight building |  |  |  |  |  |  |
|  | Mindfulness |  |  |  |  |  |  |
|  | Logical consequences |  |  |  |  |  |  |
| 5 | Praise |  |  |  |  |  |  |
|  | Modelling |  |  |  |  |  |  |
|  | Self-reward training |  |  |  |  |  |  |
|  | Stimulus control |  |  |  |  |  |  |
|  | Time out |  |  |  |  |  |  |
|  | Response cost |  |  |  |  |  |  |
|  | Response prevention |  |  |  |  |  |  |
|  | Tangible rewards |  |  |  |  |  |  |
| 6 | Relaxation training |  |  |  |  |  |  |
|  | Guided imagery |  |  |  |  |  |  |
|  | Cognitive/coping |  |  |  |  |  |  |
|  | Exposure |  |  |  |  |  |  |
| 7 | Communication skills training |  |  |  |  |  |  |
|  | Social skill training |  |  |  |  |  |  |
|  | Assertiveness training |  |  |  |  |  |  |
|  | Commands/limit setting |  |  |  |  |  |  |
| 8 | Family engagement |  |  |  |  |  |  |
|  | Family therapy |  |  |  |  |  |  |
|  | Couple/marital therapy |  |  |  |  |  |  |
| 9 | Directed play |  |  |  |  |  |  |
|  | Play therapy |  |  |  |  |  |  |
| 10 | Therapeutic letters |  |  |  |  |  |  |
| 11 | Referral to: | | | | | | |
| 12 | Other | | | | | | |

### Related Activities/Actions please tick (√) and include details as appropriate

| √ | Activity | With |
| --- | --- | --- |
|  | School Liaison |  |
|  | External Case Conferencing |  |
|  | Internal Case Conferencing |  |
|  | External Supervision/Advice |  |
|  | Client Review by Psychiatrist |  |
|  | Other Professional Liaison |  |

### Clinical incidents and medication:

Has the young person:

☐ Engaged in deliberate self-harm

☐ Been admitted to an inpatient unit

☐ Commenced a new medication

☐ Attempted suicide

☐ Been admitted to other hospital

☐ Ceased a new medication

Medication:___________________________________________________________

Dose: ________________________________________________________________

☐ Other incident:

____________________________________________________________________________________________________________________________________________________________________________________________________________________________________________________________________________________________________________

### Comments

____________________________________________________________________________________________________________________________________________________________________________________________________________________________________________________________________________________________________________________________________________________________________________________________________________________________________________________________________________________________________________

### Notes on Completion

Treatment Recording Sheet (TRS) Guide for CAMHS

Derived from the Hawaii CAMDS Service Provider Monthly Treatment and Progress Summary

**Number of Sessions**: Is the number of times you have worked with the adolescent or significant others during the preceding month

**Stage of Treatment**: Please tick where on the continuum you think you are at with this client (i.e. how close to intake versus closure)

**Intervention Strategies—ONLY** RELATING TO SINCE YOU REFERRED THEM TO THE ATTEND STUDY

Please tick/cross (x, xx, xxx according to frequency) any intervention strategies you used with this client or their significant others since you referred them to the ATTEND study. There is no limit to how many interventions may be checked. If other strategies were employed that are not included, please record the name of these in the “other” box

Definitions of Intervention Strategies

**Note:** In defining interventions the term “client” is used generically to refer to the child/adolescent, parent(s), family members, school or others.

**1.**

**Rapport Building**—Strategies used when the immediate aim is to increase the quality of the relationship between the client and the clinician. e.g. self-disclosure, reflection, play.

**Risk Assessment**—Assessment of risk of harm to self or others.

**Functional Analysis**—Arrangement of antecedents and consequences based on a functional understanding of the client’s behaviour. This goes beyond straightforward application of other behavioural techniques.  **Mental Status Exam**—Assessment of the client’s mental status (including appearance, behaviour, mood, affect, thought, speech, perceptual disturbance, insight, and judgement).

**2.**

**Motivational Interviewing**—Strategies designed to increase the client’s readiness to participate in additional therapeutic activity or programs. These can involve cost-benefit analysis, persuasion, or other approaches.

**Supportive Listening**—Reflective discussion with the client to demonstrate warmth, empathy, and positive regard without suggesting solutions or alternative interpretations.

**Emotional Processing**—Activation of emotional memories in conjunction with new and incompatible information about those memories.

**Problem Solving**—Techniques designed to bring about solutions to targeted problems, usually with the intention of imparting a skill for how to approach and solve future problems.

**Psychoeducation**—The formal review of information with the client about the development of a problem and its relation to a proposed intervention.

**Goal Setting**—Development of realistic, achievable goals to direct behaviour. Goals are identified and developed in collaboration with the client.

**Activity Scheduling**—The assignment or request that the client participate in specific activities outside of therapy time, with the goal of promoting or maintaining involvement in satisfying and enriching experiences.

**3.**

**Crisis Management**—Immediate problem solving approaches to handle urgent or dangerous events. This might involve defusing an escalating pattern of behaviour and emotions either in person or by telephone, and is typically accompanied by debriefing and follow-up.

**Maintenance/Relapse Prevention**—Exercises and training designed to consolidate skills already developed and to anticipate future challenges, with the overall goal to minimize the chance that gains will be lost in the future

**Practical Support**—Providing tangible support e.g. organizing food vouchers, arranging transport, obtaining financial assistance.

**Line of Sight Supervision**—Direct observation of the client for the purpose of assuring safe and appropriate behaviour.

**Monitoring Training**—The ongoing assessment of some target index by a significant other, or client self-monitoring of a target behaviour.

**4.**

**Interpretation**—Reflective discussion or listening exercises with the client designed to yield therapeutic interpretations.

**Catharsis**—Strategies designed to bring about the release of intense emotions, with the intent to develop mastery of affect and conflict.

**Insight Building**—Techniques designed to help the client achieve greater self-understanding.

**Mindfulness**—Techniques that facilitate present-focused, nonevaluative observation of experiences as they occur, with a strong emphasis on the here and now. This can involve the client’s conscious observation of feelings, thoughts, or situations.

**Logical Consequences**—Training others in allowing the child/adolescent to experience the negative consequences of poor decisions or unwanted behaviours, or to deliver consequences in a manner that is appropriate for the behaviour performed.

**5.**

**Praise**—Applying social reinforcements for appropriate behaviour(s)

**Modelling**—Demonstration of a desired behaviour by the clinician, peers, or others to promote the imitation and subsequent performance of that behaviour.

**Self-Reward Training**—Techniques designed to encourage the client to self-administer positive rewards contingent on performance of the target behaviour.

**Stimulus Control**—Strategies to identify specific triggers for problem behaviours and to alter or eliminate the triggers in order to reduce or eliminate the behaviour.

**Time Out**—The training of or direct use of a technique which removes the client from all reinforcement for a specified period of time following the performance of an identified, unwanted behaviour.

**Response Cost**—Training parents or teachers how to use a point or token system in which negative behaviours result in the loss of points or tokens for the child/adolescent.

**Response Prevention**—Strategies designed to prevent a maladaptive behaviour that typically occurs habitually or in response to emotional or physical discomfort.

**Tangible Rewards**—Training parents or others involved in the social ecology of the child/adolescent to administer tangible rewards to reinforce desired behaviours. Rewards can include tokens, social reinforcers, etc.

**6.**

**Relaxation**—Techniques or exercises designed to induce physical or mental relaxation. Can include breathing exercises, meditation, and visual imagery.

**Guided Imagery**—Use of visual or guided imagery techniques for the purpose of mental rehearsal of successful performance.

**Cognitive Coping**—Techniques designed to alter interpretation of events and enhance coping through examining and challenging the client’s reported thoughts, typically through the generation and rehearsal of alternative counter-statements.

**Exposure**—Techniques or exercises that involve direct or imagined experience with a target stimulus, whether performed gradually or suddenly, and with or without the clinician’s elaboration or intensification of the meaning of the stimulus.

**7.**

**Communication Skills**—Training clients to communicate more effectively with others. Can include a variety of communication strategies e.g. active listening, “I” statements.

**Social Skills Training**—Providing information and feedback to improve interpersonal verbal and non-verbal functioning. Can include rehearsal, role play and in vivo exercises.

**Assertiveness Training**—Exercises or techniques designed to promote the client’s ability to be assertive with others, usually involving rehearsal of assertive interactions.

**Commands/Limit Setting**—Training significant others in how to give directions and commands and set limits in such a manner as to increase client compliance.

**8.**

**Family Engagement**—The use of strategies to facilitate the family’s positive interest in participation in an intervention.

**Family Therapy**—A set of family-centred approaches designed to shift patterns of relationships and interactions within a family.

**Couple/Marital Therapy**—Techniques used to improve the quality of the relationship between caregivers.

**9.**

**Directed Play**—Exercises involving the child/adolescent and a significant other playing together in a specific manner to facilitate improved verbal communication and nonverbal interaction.

**Play Therapy**—The use of play as a primary therapeutic strategy. May include the use of play for clinical assessments or clinical interpretations. Differs from Directed Play where the specific focus is on facilitating communication. Play therapy can be directive (therapist leads the way) or non-directive (child decides what to do within safe boundaries).

**10.**

**Medication/Pharmacotherapy**—Prescription, monitoring or review of medication(s) to manage emotional, behavioural, or psychiatric symptoms.

**Therapeutic Letters**—Sending a letter to the client between sessions to initiate engagement, offer support or facilitate positive change.

**11.**

**Referral**—Record any referrals for services or supports.

**12.**

**Other**—Record any other interventions you have used. Include a short description of the strategies used.

Critical Incidents and Medication

Record any critical incidents that have occurred since you referred them to the ATTEND study and changes made to any medication (e.g. medication added or ceased or dose change).

Thank you!!
