## Appendix A Topic Guides for "Mental health pathways and treatment as usual for young people experiencing depression in the United Kingdom: A mixed methods study"

**Topic Guide 1: For interviewing senior clinicians**

An interview guide for senior clinicians (3-5 per site) to establish the care pathway/s for young people with depression, and to explore the fit of the Treatment Recording Sheet (TRS) with that pathway ahead of its use in the site.

Time allowed: 30 minutes

**Aims of the study**

Before we can test whether MAC works and is value for money, we need to answer four questions.

1. **Who should be trained to deliver mindfulness, and how should they be trained?**
2. **What does 'usual' treatment look like for this group of young people?**
3. **How can young people and their carers best be recruited to a study on mindfulness?**
4. **We want to know if people would be willing to provide blood and saliva samples**

We will use what we have learned to plan a future programme of work that will definitively test whether MAC works, is value for money, whom might it work best for and how best can we identify, train and support clinicians to deliver MAC.

**Introduction**

This interview will contribute to addressing the second question, ‘what does 'usual' treatment look like for this group of young people?’ This will help us understand how young people in this group are currently treated by CAMHS, to ensure we target those young people most likely to benefit from mindfulness and plan a larger trial that has the most appropriate comparator.

Interviewer to explain:

Tape/audio-recording

Confidentiality

*You’ve been selected for an interview because you are a senior clinician working in one of our MAC intervention sites. We’d like to ask you some questions about the care pathways in your service for young people with depression and your thoughts on the use of a Treatment Recording Sheet (TRS) to record therapy or intervention provided to young people in your service.*

*Do you consent to being interviewed (will have received and completed consent form beforehand, this is to confirm consent) and can we record you?*

1. **Can you briefly describe your role in your service?**
2. **Within your service can you describe the care pathway or pathways for a young person (YP) with depression?**

Prompts:

- How do they get referred to CAMHS or enter the service?
- What kind of assessment takes place after referral? Are any YP discharged at this point? Are they different for YP who have been re-referred after an earlier discharge?
- Would the assessments made be different if the referral was classified as urgent?
- Does your service have a particular pathway for children with depression?
- Is this pathway specific to depression or for emotional disorders more generally?
- Are YP with depression likely to be anywhere other than a Mood Pathway in your system? Check what happens if comorbid condition, such as ASC or eating disorders
- What are the first-line treatments available to them? Do these alter depending on the pathway a patient was referred via?
- For how long and at what frequency are these treatments offered?
- Is there a typical timeline for YP in this pathway? If no, what are the key factors for increasing length of time in this pathway?
- How long is the average waiting time between referral to the service and treatment starting? What are the longest and quickest times?
- Are YP offered any form of therapy or support if they need to wait for a first-line treatment?
- What does care look like between diagnosis and receiving a first-line intervention/treatment?

3. **When patients relapse or do not recover after first line treatments, can you describe the pathway for these individuals?**

Prompts:

- What assessment determines that a second-line treatment is needed?
- What is the timeline for receiving a second-line treatment?
- What are the second-line treatments available to YP with depression?
- Are YP offered any form of therapy or support if they need to wait for a first-line treatment?
- Do you offer any form of mindfulness-based cognitive therapy?
- How do you know/assess whether a YP is ready for discharge from the service?

We are exploring the use of a Treatment Recording Sheet for use in the main trial of MAC. This would require the lead clinician for every YP involved in the trial to record any non-specified therapy or psychological intervention provided to young people and /or parents / families (e.g. goal setting, supportive listening, family therapy) and the frequency with which they are provided at each data collection point. This will help us to compare treatment strategies across sites.

4. **What do you think about the TRS and its relevance for your service care pathway?**
a. If this is not something you’ve come across before, what are your first impressions of the TRS?
b. If you have used this before, in what capacity and what was your experience?

Prompts:

- Positive or negative aspects
- Are there any changes you would make to the TRS to make it more acceptable / usable in your service context?
- Do you feel there are better ways to capture this information? Examples?

5. **Do you feel it is feasible to complete this at each data collection point?**

Prompts:

- Why do you feel it is not feasible? What is it about the TRS specifically or about the frequency of completion?
- If not feasible, what could be changed to make it more feasible in your service context?

6. **Is there anything else you want to tell us about the care pathway/s for YP with depression in your service?**

Would you be happy for us to follow up with you with any uncertainties we discover about these pathways/to sense check a flowchart?

**Topic Guide 2: Case-managing clinicians**

An interview guide for case-managing clinicians (5-10 per site) to establish what the YP in both the MAC arm and Treatment As Usual (TAU) arm received.

Time allowed: 30 minutes

**Aims of the study:**

Before we can test whether MAC works and is value for money, we need to answer four questions.

5. **Who should be trained to deliver mindfulness, and how should they be trained?**
6. **What does 'usual' treatment look like for this group of young people?**
7. **How can young people and their carers best be recruited to a study on mindfulness?**
8. **We want to know if people would be willing to provide blood and saliva samples**

We will use what we have learned to plan a future programme of work that will definitively test whether MAC works, is value for money, whom might it work best for and how best can we identify, train and support clinicians to deliver MAC.

**Introduction**

This interview will contribute to addressing the second question, ‘what does 'usual' treatment look like for this group of young people?’ This will help us understand how young people in this group are currently treated by CAMHS, to ensure we target those young people most likely to benefit from mindfulness and plan a larger trial that has the most appropriate comparator.

Interviewer to explain:

Tape/audio-recording

Confidentiality

*You’ve been selected for an interview because you are a clinician involved in the ATTEND study. We’d like to ask you some questions about the care pathways in your service for young people with depression to establish what the YP in both the MAC arm and TAU arm actually received.*

*Do you consent to being interviewed (will have received and completed consent form beforehand, this is to confirm consent) and can we record you?*

1. **Can you briefly describe your role in your service and your involvement in the ATTEND study?**
2. **Can you describe the typical care pathway for YP who were allocated to receive the MAC intervention?**

Prompts:

- Were they in the ‘mood pathway’ in your system or did they come from elsewhere?
- What first-line treatments had they received prior to MAC?
- Why/how were they assessed as requiring a second-line treatment?

3. **Did YP allocated to the MAC group receive any additional treatment/s or support beside MAC?**

Prompts:

- What other treatments were considered for the YP?
- If nothing additional, was this the same for all YP involved in MAC?
- If yes, what treatment/s did they receive / for how long / at what intensity typically?
- To what extent (in terms of time) did it overlap with their engagement in MAC?
- What did this treatment aim to achieve / what was it targeting? Is this separate or similar to the aims of MAC?
- Did their parents/carers or family receive any additional support or service in relation to the YP’s mental health?

4. **Can you describe the care pathway/s for those YP who received TAU?**

Prompts:

- What first-line treatments had they received, if different to YP who received MAC?
- What second-line treatments were they offered? And why?
- What was the timeline for receiving this treatment?
- Did the YP or their carers receive any mindfulness-based cognitive therapy, or similar?

5. **What was your experience of using the Treatment Recording Sheet (TRS) for the ATTEND study?**

Prompts:

- Positive or negative aspects
- Are there any changes you would make to the TRS to make it more acceptable / usable in your service context?
- Do you find it feasible to complete at each data collection point?
- If no, why do you feel it is not feasible? What is it about the TRS specifically or about the frequency of completion?

**Topic Guide 3: For interviewing families**

An interview guide for families (10-15 per site) to understand their experience of the MAC intervention or TAU arm.

Time allowed: 1 hour per dyad

*Note – we will invite YP and their carers as dyads but either may decide to be interviewed separately if preferred*

**Aims of the study**

Before we can test whether MAC works and is value for money, we need to answer four questions.

1. **Who should be trained to deliver mindfulness, and how should they be trained?**
2. **What does 'usual' treatment look like for this group of young people?**
3. **How can young people and their carers best be recruited to a study on mindfulness?**
4. **We want to know if people would be willing to provide blood and saliva samples**

We will use what we have learned to plan a future programme of work that will definitively test whether MAC works, is value for money, whom might it work best for and how best can we identify, train and support clinicians to deliver MAC.

**Introduction**

This interview will contribute to addressing the second question, ‘what does 'usual' treatment look like for this group of young people?’ This will help us understand how young people in this group are currently treated by CAMHS, to ensure we target those young people most likely to benefit from mindfulness and plan a larger trial that has the most appropriate comparator.

Interviewer to explain:

Tape/audio-recording

Confidentiality

*You’ve been selected for an interview because you are a family / young person / carer in one or our MAC intervention sites and you consented to take part in the ATTEND study. We’d like to ask you some questions about the MAC treatment you have recently taken part in (if you received this), and the support you’ve received from your CAMHS service, and also any feedback you have about your involvement in the ATTEND study.*

*Do you consent to being interviewed (will have received and completed consent form beforehand, this is to confirm consent) and can we record you?*

1. **Could you start by telling us a bit about your care to date and what treatment you had received prior to being referred to the ATTEND study?**

Prompts:

- This can include input from primary care, education and community settings
- Who suggested you went to CAMHS? Did they refer you?
- If you can remember, how long did you wait from referral to your first appointment?
- How did CAMHS explain to you the struggles you were having? (did they offer a diagnosis)
- If you can remember, how long was it between the decision for treatment and you getting treatment?
- The length and frequency of treatment the YP had received prior to the study starting
- What was your experience of that treatment (positive/negative, perception of progress/no progress)
- What’s happened since the ATTEND study started in December 2020/January 2021? What has the process been for you?

2. **What treatment (therapy or medication) have you been offered and what have you received since being involved in the ATTEND study?**

Prompts:

- Treatment, duration/frequency, time to receive any subsequent treatment
- If you’ve received more than one treatment, could you tell us what?
- If MAC intervention arm YP, have you received anything besides MAC?

IF THIS IS A CONTROL/TAU PARTICIPANT THEN SKIP TO QUESTION 7

3. **(Both Arms) Did you know much about mindfulness-based cognitive therapy before you agreed to take part in the ATTEND study?**

Prompts:

- What did you know about it already? What would it have been helpful to know?
- What were your expectations going in? Was it how you expected it might be?

4. **(Intervention Arm only) What was your experience of MAC (the treatment)?**

Prompts:

- Did you find it helpful? Did you enjoy it or get something out of the experience of attending?
- How did you find the online delivery of the course? Would you have preferred the group to be face-to-face?
- Is there anything that might have made the experience better? Is there anything you would change about it?

5. **(Intervention Arm only) Has anything from the mindfulness course played a part in staying well during the trial? If so, can you describe how?**

Prompts:

- Were there any techniques, ideas, a response plan that made a difference?
- Why/why not?
- How did they help?
- When did they help (i.e. straight away or only after a few weeks)?

6. **(Intervention Arm only) How many days per week do you practice (either formally or informally)? How long are those practices?**
   a. Prompts: Is there any additional information or support that would have aided your practice at home? Were the materials from the programme useful for your home practice?
7. **(TAU Arm only) What was your experience of the therapy or treatments you received from the service?**

Prompts:

- Did you find it/them helpful?
- Have any of these treatments been delivered digitally/online? If so, do you prefer online or face-to-face formats?
- Is there anything that might have made the experience better? Is there anything you would change about it?

8. **Do you think you’ve made progress or changed during the time of the study?**

Prompts:

- If yes, how do you think you’ve progressed or changed? In what way? Why do you think that is?
- What things have you noticed that show you have progressed or changed?
- How do you assess that change / how do you notice it? (E.g. when you do certain things, how you feel in particular situations etc.)
- If no, why do you think that might be? What do you think would need to be different for you to progress/change? (E.g. more time / different treatment / not able to assess change)

9. **Reflecting on the whole process and your involvement with the ATTEND study, how would you describe your experience?**

Prompts:

- What could have been done better?
- Do you think if we repeated the study there are better ways we could communicate the random allocation of YP to receive an intervention being tested (like MAC) or to continue with services as usual? Do you think YP will mind which group (Treatment or TAU) they get assigned to?
- How was your experience of the research process? Is there anything we could change to make the process easier to understand or contribute to?

10. **In future research trials of the MAC intervention, we are interested in both biological and psychological mechanisms. How would you feel about providing blood or saliva samples for this type of study?**

Prompts:

- How willing would you be to do that?
- What barriers or challenges might this involve for you? Do you think it would be acceptable for other families/YP? If no/yes – why do you think that?

What could be done to make this more acceptable/less challenging
